# Gastric emptying and nutrient absorption of pea protein products differing in heat treatment and texture: a randomized *in vivo* crossover trial and *in vitro* digestion study

**DOI:** 10.1101/2023.09.13.23295474

**Authors:** Julia J.M. Roelofs, Elise J.M. van Eijnatten, Patteela Prathumars, Joris de Jong, Ron Wehrens, Diederik Esser, Anja E.M. Janssen, Paul A.M. Smeets

**Author notes:** Pubmed indexing: Roelofs JJM, van Eijnatten EJM, Prathumars P, de Jong J, Wehrens R, Esser D, Janssen AEM, Smeets PAM. Source of support: Wageningen University and Research. Corresponding Author Name: Julia Roelofs Mailing address: Stippeneng 4, 6708 WE Wageningen. Ethical statement and registration: "Ethical approval for the involvement of human subjects in this study was granted by the Medical Ethical Committee of Wageningen University (Reference number: NL74440.081.20, dated April 13^th^, 2021) and is in accordance with the Helsinki Declaration of 1975 as revised in 2013. This study was registered with the Dutch Trial Registry under number NL9413 (record can be retrieved from the International Clinical Trials Registry Platform at https://trialsearch.who.int/Trial2.aspx?TrialID=NL9413).

## Abstract

**Background:** Pea protein is an interesting alternative for animal-based proteins due to its good availability, low cost and relatively balanced amino acid (AA) profile. Its digestibility may be affected by heat treatment and food texture.

**Objectives:** To study *in-vivo* AA absorption kinetics and gastric behavior of pea protein products differing in heat treatment and texture and compare this with *in-vitro* digestion.

**Design:** Fourteen males participated in a randomized crossover trial. Iso-caloric and iso-volumetric treatments were a 420-mL heated drink, 420-mL unheated drink and 105-g heated gel (semi-solid) consumed with 315 mL water, all containing 20 g pea protein. Gastric MRI scans were made until 90 minutes post-prandial. Blood samples were collected at baseline and up to five hours. All treatments were tested with an *in-vitro* digestion model (INFOGEST).

**Results:** Heat treatment did not alter AA absorption kinetics and gastric emptying. Time to maximum peak was delayed for the gel treatment (total AAs: 66.9 versus 48.0 min for both drinks, essential AAs: 75.4 versus 50.0 and 46.6 min for the drinks). For the gel treatment initial emptying was faster due to the rapid passage of water. *In-vitro*, the degree of hydrolysis was highest for the unheated drink in the gastric phase and for the gel treatment in the intestinal phase.

**Conclusion:** Heat treating pea protein products does not affect digestion. In contrast, texture of pea protein products can be altered to influence the rate of gastric emptying and AA absorption without affecting total AA absorption.

## 1. Introduction

Protein is an essential building block for the growth and renewal of tissue (Atherton & Smith, 2012). For certain populations, such as older adults, athletes and critically ill, it can be difficult to obtain the necessary amount of protein from the diet (Coelho-Júnior, Rodrigues, Uchida, & Marzetti, 2018; Liao, et al., 2017; Sieber, 2019). It is therefore important that the protein we ingest is properly digested and absorbed, so that it can be used for protein synthesis (Fardet, Dupont, Rioux, & Turgeon, 2019; Mahe, et al., 1996; van Vliet, Burd, & van Loon, 2015). Digestion consists of a series of mechanical, physiological, and biochemical processing steps leading to the breakdown of food structures that eventually allows for absorption and utilization of nutrients (Mackie, 2019). Despite all these processing steps, some proteins are still poorly digested. This is especially the case for plant-based proteins, which often have a lower digestibility (Berrazaga, Micard, Gueugneau, & Walrand, 2019; Pasiakos, Agarwal, Lieberman, & Fulgoni, 2015). However, due to the growing population worldwide, animal-based protein puts a strain on the environment (Katz, 2019). Because of this, the demand for more sustainable plant-based proteins is rapidly growing. Therefore, it is important to explore how the digestibility of plant-based proteins may be improved.

The lower digestibility of proteins from plants is explained by the intact cell wall that hinders direct contact between intracellular macronutrients and the digestive enzymes. This slows down or even completely prevents the access of proteases to the cell contents and limits intracellular protein hydrolysis. Thus, the digestibility of plant-based proteins depends on the fraction of broken cells generated during their processing (Zahir, Fogliano, & Capuano, 2018). Food processing such as the isolation of proteins, alters the chemical and physical characteristics and can increase the nutritional value of food products (Joye, 2019). Plants also contain anti-nutritional factors. These are compounds that reduce nutrient utilization and/or food intake of plants or plant products used as human foods which can be removed or inactivated by processing (Thakur, Sharma, & Thakur, 2019). In addition, the quality of plant-based protein is often lower compared to animal-based protein. Animal-based protein has the highest protein quality as determined by the Digestible Indispensable Amino Acid Score (DIAAS). The DIAAS of animal-based proteins is typically greater than 100, indicating excellent quality, while for plant-based proteins it is generally below 75, indicating lower quality (Herreman, Nommensen, Pennings, & Laus, 2020).

Several plant-based proteins, from crops such as wheat, soy and pea, are increasingly used in foods. With its good availability, low cost and relatively good quality for a plant-base protein (DIAAS = 70), pea protein is one of the better alternatives for animal-based proteins in functional food applications (Bailey, Fanelli, & Stein, 2023; Lu, He, Zhang, & Bing, 2020). Although there is ample information about the digestion of traditional protein sources, the digestion of pea protein and the influence of intensive processing on its digestion is not known in detail (Rivera del Rio, et al., 2020). This is essential to evaluate its potential as a nutritious sustainable protein source.

Digestion of food products is predominantly studied with *in vitro* digestion models (Muttakin, Moxon, & Gouseti, 2019). Although these models are based on *in vivo* data, they obviously do not account for all factors, such as the mixing of the food in the stomach. Therefore, *in vivo* research is needed to understand to what extent *in vitro* models represent *in vivo* digestion. Magnetic Resonance Imaging (MRI) allows for visualization and quantification of gastric processes such as gastric emptying, emulsion stability and coagulation (Smeets, Deng, van Eijnatten, & Mayar, 2021). In addition, measuring AA concentrations provides information on differences in absorption kinetics. Although it is not possible to directly relate gastric emptying with subsequent AA absorption because of all intermediate processes involved, combining these measurements does provide more insight in the overall differences between products.

Gastric emptying is largely determined by the chemical characteristics of food, such as the energy density and macronutrient content, but also by physical characteristics, such as texture (Camps, Mars, De Graaf, & Smeets, 2016; Marciani, et al., 2001; Roy, et al., 2022). The food matrix plays an important role in digestibility because of its influence on the kinetics of transit and hydrolysis of macronutrients. For example, liquids empty faster from the stomach compared to semi-solid foods (Camps, et al., 2016; Clegg & Shafat, 2014; Mackie, Rafiee, Malcolm, Salt, & van Aken, 2013; Zhu, Hsu, & Hollis, 2013).

The isolation of plant-based proteins often includes a thermal denaturation step. Thermal denaturation of proteins may either improve of decrease their digestibility, depending on the type of protein and severity of the heat treatment. Proteins either lose their tightly folded structure, resulting in a higher accessibility of the peptide chain for enzymes, or they will aggregate, thereby impairing digestion (Joye, 2019). *In vitro* work on pea protein showed that heating disrupts the structure, thereby increasing the number of smaller better digestible particles. Conversely, these heat-induced aggregates are up to 50% less digestible compared to before the heat treatment (Mulet-Cabero, Mackie, Wilde, Fenelon, & Brodkorb, 2019; Rivera del Rio, et al., 2020).

The aim of this study was to measure *in vivo* AA absorption kinetics and gastric behavior of pea protein products differing in heat treatment and texture. In addition, we aimed to compare *in vitro* digestion data with the *in vivo* data.

## 2. Methods

### 2.1. In vivo trial

#### 2.1.1. Design

The study was a randomized crossover trial in which healthy men underwent gastric MRI scans and blood sampling before and after consumption of three pea protein products. Primary outcomes were plasma AA absorption kinetics and gastric volume over time. Secondary outcomes included MRI markers of digestion (T_2_, MT), plasma glucose and insulin concentrations and appetite and nausea ratings (hunger, fullness, thirst, desire to eat, prospective consumption and nausea). The procedures followed were approved by the Medical Ethical Committee of Wageningen University in accordance with the Helsinki Declaration of 1975 as revised in 2013. This study was registered with the Dutch Trial Registry under number NL9413. The record can be retrieved from the International Clinical Trials Registry Platform at https://trialsearch.who.int/Trial2.aspx?TrialID=NL9413. All participants signed informed consent.

#### 2.1.2. Participants

Healthy (self-reported) males aged 18-55 y and with a BMI between 18.5-25.0 kg/m^2^ were included. Participants were excluded if they reported a pea allergy, gastric disorders or regular gastric complaints, used medication that affects gastric behavior, used recreational drugs within 1 month prior to the study screening day, smoked more than 2 cigarettes per week, had an alcohol intake >14 glasses per week, or had a contra-indication to MRI scanning (including but not limited to pacemakers and defibrillators, ferromagnetic implants and claustrophobia). Since female sex hormones are known to influence gastrointestinal function, only males were included in the study (Gonenne, et al., 2006; Lajterer, Levi, & Lesmes, 2022; Soldin & Mattison, 2009). Participants were recruited via digital advertisements (e-mail and social media).

#### 2.1.3. Sample size

A priori sample size was estimated for both primary outcomes, i.e. AA absorption kinetics and gastric volume over time. The estimation for postprandial AA was based on the peak value and the total free AA assessed in the serum after consumption of protein products. For the peak value, a difference of 100 µg/mL was regarded as relevant with an individual difference in peak values of 100 µg/mL (Farnfield, Trenerry, Carey, & Cameron-Smith, 2009; He, Spelbrink, Witteman, & Giuseppin, 2013). Given an α of 0.05 and a power of 0.9, we estimated a requirement of 11 participants.

For gastric emptying the sample size estimation was based on gastric emptying half times of liquids from Camps, et al. (2016), and gels from Hoad, et al. (2009) taking into account intake volume and caloric content. We estimated 10 min as the minimum detectable difference which is physiologically relevant, and an average SD of 11 minutes. With a two-sided test, an α of 0.05 and a power of 0.9, this resulted in a minimum of 12 participants. To accommodate drop-out, we aimed to include 14 participants. The calculation were done using software from: http://hedwig.mgh.harvard.edu/sample_size/js/js_crossover_quant.html.

#### 2.1.4. Treatments

The three treatments were a 420-mL unheated pea protein drink, 420-mL heated pea protein drink and 105 g heated semi-solid pea protein food (gel) consumed with 315 mL water (Table 1). All treatments contained 20 g of pea protein isolate (Nutralys® F85M, Roquette, France) and were iso-caloric (153 kcal) and iso-volumetric (420 mL). In addition to pea protein isolate and water, the test foods contained vanilla aroma, chocolate aroma, cocoa powder and sweetener (See **Supplement** for exact product preparation). The heated treatments were heated in a steam oven at 90 °C for 30 minutes.

**Table 1.**
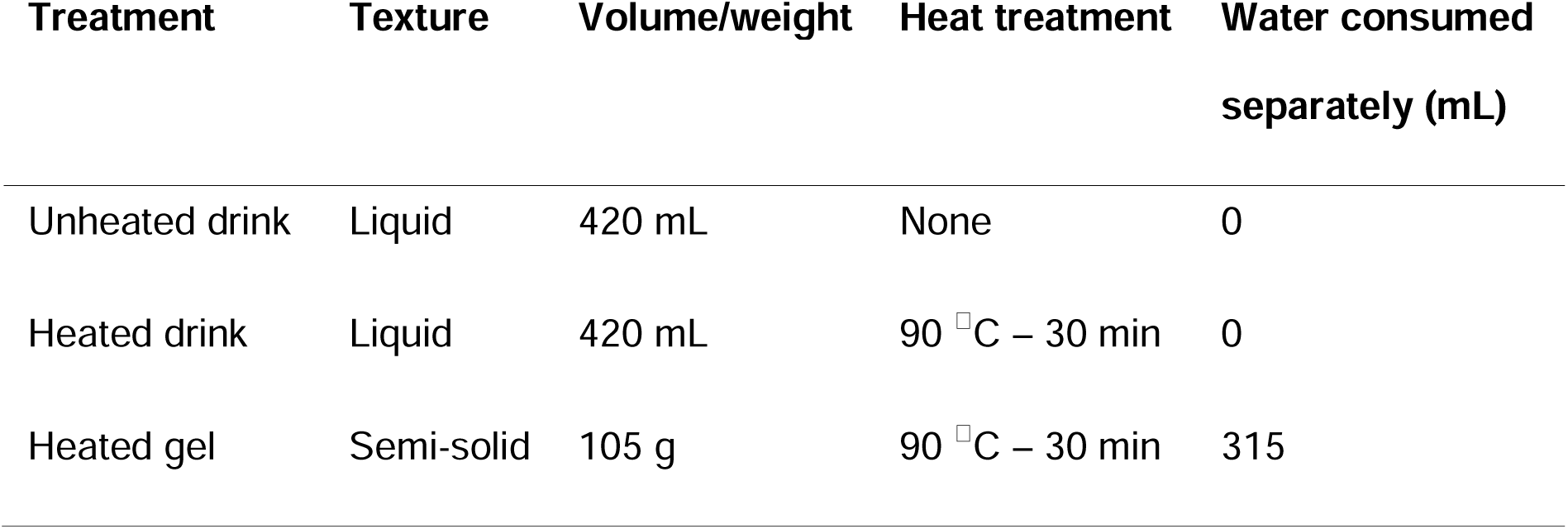
Treatment overview.

| Treatment | Texture | Volume/weight | Heat treatment | Water consumed<br>separately (mL) |
| --- | --- | --- | --- | --- |
| Unheated drink | Liquid | 420 mL | None | 0 |
| Heated drink | Liquid | 420 mL | 90 °C – 30 min | 0 |
| Heated gel | Semi-solid | 105 g | 90 °C – 30 min | 315 |

#### 2.1.5. Study procedures

The evening before the test day participants consumed a standard pasta meal (Iglo Green Cuisine Linguine Bolognese) after which their overnight fast started. During the fasting period of at least 12 hours, participants were allowed to drink water and herbal tea up to 1.5 hours prior to their visit. Upon arrival at Hospital Gelderse Vallei (Ede), a cannula was placed, a baseline MRI scan was performed, appetite and nausea ratings were taken and a blood sample was drawn. Subsequently, participants consumed one of the three treatments. For the drink, the participants were instructed to consume it over a period of 5 minutes through a straw to ensure an eating time comparable to that of the gel (mean ingestion time was 4.7 ± 0.6 and 4.7 ± 0.9 min for the heated and unheated drink respectively). For the gel treatment, participants were instructed to consume it within 10 minutes and alternate eating and drinking (mean ingestion time 6.7 ± 1.3 min). Subsequently, gastric MRI scans were performed at t = 10, 15, 20, 30, 40, 50, 60, 70, 80 and 90 minutes after the start of ingestion. Blood samples were taken at t = 30, 60, 75, 90, 120, 150, 180, 240 and 300 min. In addition, participants verbally rated their appetite and nausea on a scale from 0 to 100 every 10 minutes, up to 90 minutes (Noble, et al., 2005) (**Figure 1**).

**Figure 1.**
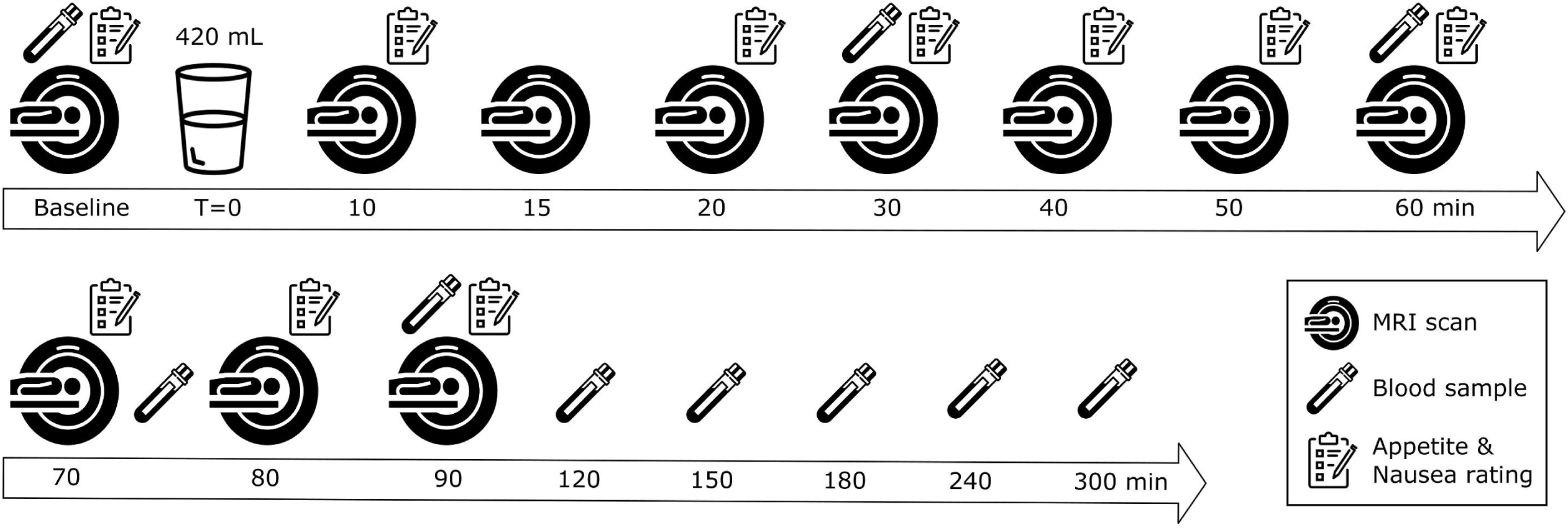
Overview of a test session.

#### 2.1.6. MRI

Participants were scanned in a supine position with the use of a 3-Tesla Philips Ingenia Elition X MRI scanner (Philips, Eindhoven, The Netherlands). A 2-D Turbo Spin Echo sequence (37 4-mm slices, 1.4 mm gap, 1 x 1 mm in-plane resolution, TR: 550 ms, TE 80 ms, flip angle: 90 degrees) was used with breath hold command on expiration to fixate the position of the diaphragm and the stomach. The scan lasted approximately 20 seconds.

Total gastric content was manually delineated on every slice by two researchers with the use of the program MIPAV (Medical Image Processing, Analysis and Visualization Version 7.4.0, 2016) (**Supplementary Figure 1**). When the volumes differed by more than 10% between the two researchers, the segmentation was re-evaluated to reach consensus. Total gastric volume for each time point was calculated by multiplying the number stomach content voxels with voxel volume, taking into account slice thickness and gap distance. The gastric volumes of the two researchers were averaged.

For the gel treatment, volumes of liquid and semi-solid content of the stomach were estimated based on voxel intensity using thresholding (Reddy & Reddi, 2017). The baseline scan was used to calculate the cut-off value for each participant. The cut-off value that was chosen included the 10% voxels with the lowest intensity, since this resulted, on average, in a volume for the semi-solid content at t = 10 minutes close to the volume of ingested (mean solid volume of 102.6 mL, SD = 17.5). This cut-off value was used for all scans in that scan session. An example of a stomach with its semi-solid and solid content marked in red can be found in **Supplementary Figure 2**.

As an approach to quantify gastric coagulation, image texture metrics of the stomach content were calculated with the use of the LIFEx software (version 7.2.10) (Nioche, et al., 2018). These image metrics provide information on the spatial patterns of voxel intensity (Thomas, et al., 2019). Four metrics were calculated: busyness, coarseness, contrast and homogeneity. Neighborhood Grey-level Difference Matrix (NGLDM) difference of grey-levels between one voxel and its 26 neighbors in 8 dimensions was used for busyness, coarseness and contrast. Busyness translates to the spatial frequency of changes in intensity. Coarseness translates to the spatial rate of change in intensity. Contrast is the local variation in grey level. The Gray-Level Co-occurrence Matrix (GLCM) method was used for homogeneity and reflects the differences in voxel intensity between the neighboring voxels. The number of grey levels for texture metric calculation was set at 64, intensity rescaling relative (ROI: min/max) and dimension processing 2D.

These texture metrics were calculated for each slice. Subsequently, a weighted mean was calculated based on gastric volume in each slice, i.e. small stomach volume areas will contribute less to the mean compared to larger areas. In the context of this paper we interpret changes in image texture metrics as reflecting changes in the degree of coagulation. An example of two stomachs with their corresponding image texture measures indicating relatively high and low coagulation can be found in **Supplementary figure 3.**

The gel treatment was not considered for this analysis, since the presence of dark gel particles in the stomach would yield very different image texture metrics than those of the two drinks. In addition, the analysis was only performed for the postprandial scans. Due to the exploratory nature of these measures, we did not correct for multiple testing.

#### 2.1.7. Clinical chemistry

Blood samples were drawn from the cannula into sodium-fluoride (3 ml) and EDTA (4 ml) tubes. After collection, sodium-fluoride tubes were centrifuged at 1000 g for 10 min at 22°C, to obtain blood plasma. The EDTA tubes were centrifuged at 1000 g for 10 min at 4°C. Following centrifugation, aliquots of 250 µl and 500 µl were pipetted in 2.0 ml cryo-vials and 5 ml tubes and stored at -80°C until they were analyzed in bulk.

Free AA concentrations were analysed as described previously (Mes, et al., 2022) and based on the Waters AccQ Tag method for AA analysis. To determine glucose concentrations, the plasma samples were processed using the Atellica CH Glucose Hexokinase_3 (GluH_3) assay kit and quantified using the Atellica CH analyzer (Siemens Healthineers, Netherlands) by a hospital laboratory (Ziekenhuis Gelderse Vallei, Ede, The Netherlands). The lower detection limit was 0.2 mmol/l and interassay CVs were at most 4.5%. The plasma samples were processed and quantified using an enzymatic immunoassay kit (ELISA, Mercodia AB, Sweden) to determine insulin concentrations. The lower detection limit was 6 pmol/l and inter-assay CVs ranged between 0.3 and 20.0%.

### 2.2. In vitro digestion

A static *in vitro* digestion was performed using the INFOGEST digestion protocol for all three treatments (Brodkorb, et al., 2019). Gastric digestion was performed for 2 hours followed by 2 hours of intestinal digestion. The degree of hydrolysis and size distribution of the soluble peptides were measured at 30-minute intervals in the gastric phase and at 60-minute intervals in the intestinal phase. Moreover, the unheated and heated drink were tested in a semi-dynamic system. The complete protocol can be found in the **Supplement**.

To measure the particle size of the precipitation in the drinks, a mastersizer was used with an obscuration limit of 4-20%, a reflective index of 1.46 and absorption of 0.1. Non-spherical particle size was selected. The samples were measured at 0, 60 and 120 minutes after the start of gastric digestion. Cocoa powder was tested separately to check for any influences on the measurements of the drinks. Results are reported as volume density.

### 2.3. Statistical analysis

AA concentrations over time were analyzed using the software described in Wehrens (Submitted for publication, 2023). In short, peak heights, time to maximum peak and area under the curve of serum AA were calculated for total AAs (TAA) and essential AAs (EAA). For these three parameters of interest, a linear mixed model was used to assess differences between treatments. Analysis was performed in R version 4.1.3.

Further analyses were performed in R statistical software (version 4.0.2). Differences in gastric content volume over time were tested with the use of linear mixed models, testing for main effects of time, treatment and treatment*time interactions, with baseline gastric volume as a covariate. Tukey HSD-corrected post-hoc tests were used to compare individual time points. AUC of gastric content volume over time was calculated using the trapezoidal rule. Differences in AUC between treatments were tested by using one-way repeated measures ANOVA.

Differences in the texture metrics (busyness, coarseness, contrast and homogeneity) of the postprandial gastric volume over time were tested using linear mixed models, with time, treatment and treatment*time as fixed factors. Tukey HSD corrected post-hoc tests were used to compare individual time points.

Differences in glucose concentrations, insulin concentrations and appetite and nausea ratings over time were tested by using linear mixed models, testing for main effects of time, treatment and treatment*time interactions. Baseline values were added as covariate. Tukey HSD-corrected post-hoc tests were used to compare individual time points.

For each variable, normality of the data was confirmed with quantile-quantile (Q-Q) plots of the residuals. For insulin, contrast and nausea a logarithmic transformation was applied to create a normal distribution. The significance threshold was set at p = 0.05. Data are expressed as mean ± SD unless stated otherwise.

## 3. Results

In total, 14 men participated in the study (age: 23.0 ± 3.8 y, BMI: 22.2 ± 1.7 kg/m^2^). Two participants dropped out after one test session. Hence, an additional two participants were recruited. Two participants completed only two test sessions due to Covid-19 infection related quarantine (**Supplementary Figure 4**).

### 3.1. Blood amino acid kinetics

**Figure 2** shows the curves of TAA and EAA over time. AUC did not differ between the three treatments (**Figure 3 and Supplementary Figure 5**). Individual curves of TAA and EAA can be found in **Supplementary Figure 6**. For the individual AA, only tyrosine showed a significant lower AUC for the gel treatment compared to unheated and heated drink (234 µM*min (CI: 201 – 268) compared to 304 µM*min (CI: 249 – 359) and 311 µM*min (CI: 262 – 360) respectively). AUCs of the other AAs did not differ significantly. Curves of the individual AAs are shown in **Supplementary Figure 7**.

**Figure 2.**
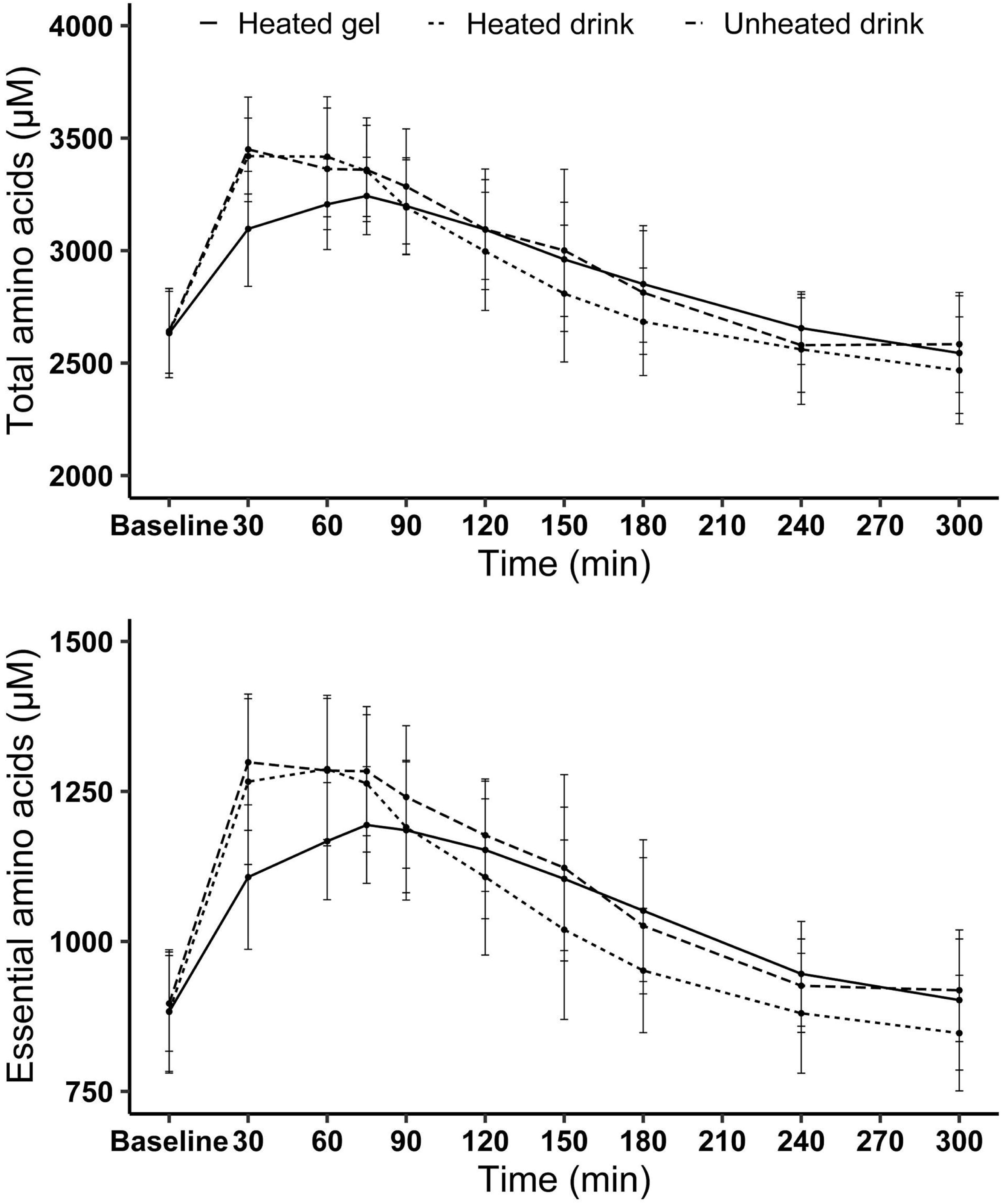
Total amino acids (top) and essential amino acid (bottom) levels over time after consumption of the three pea protein products (mean ± SD).

**Figure 3.**
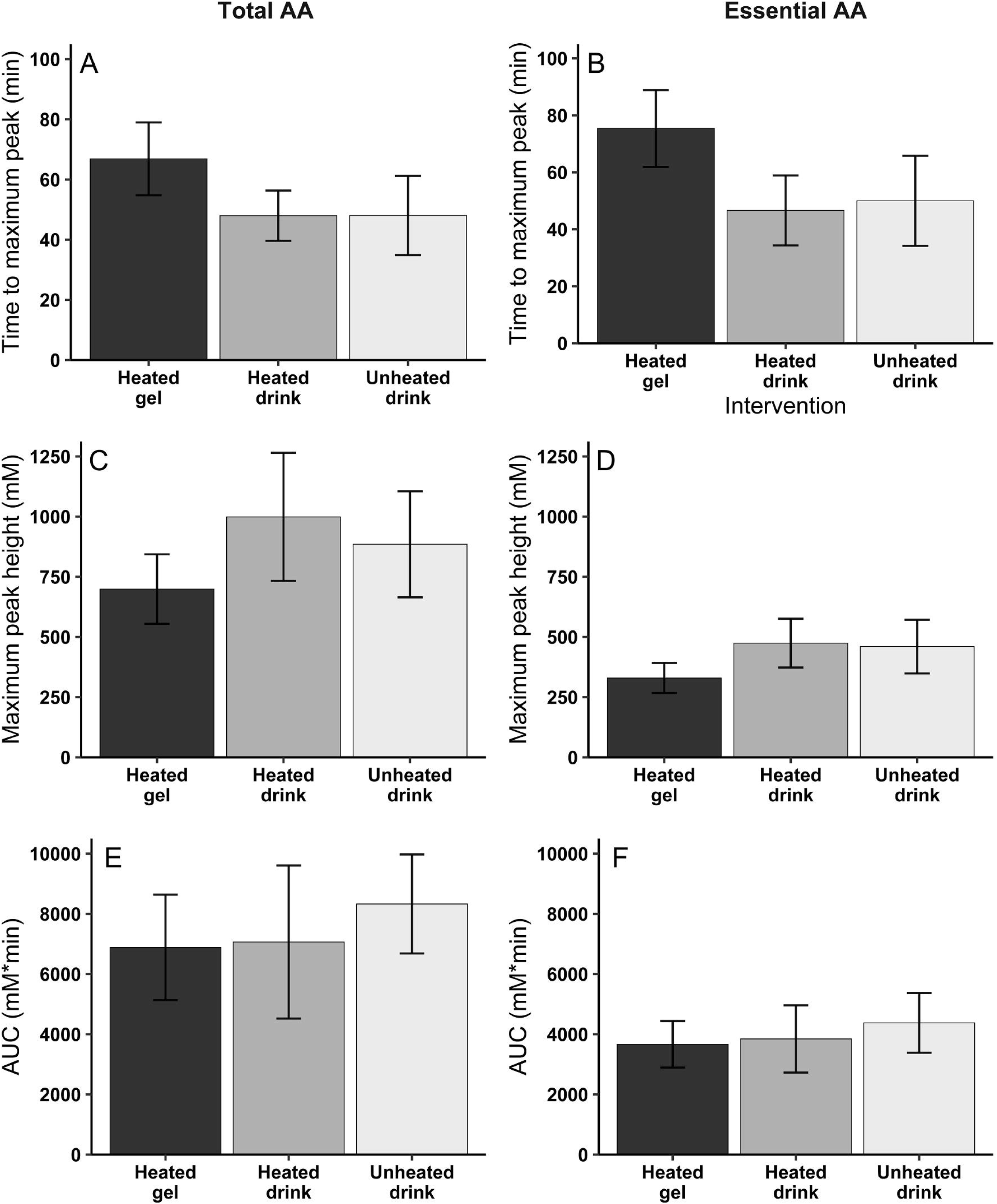
Mean ± SD time to maximum peak (min) (A and B), maximum peak height (µM) (C and D) and AUC (mM*min) (E and F) of the three pea protein products for total AA (left panel) and essential AA (right panel).

Maximum peak height for EAA was 131 and 145 µM lower for the gel treatment compared to the unheated and heated drinks (330 µM (CI: 290 – 369) compared to 460 µM (CI: 389 – 531) and 475 µM (CI: 407 – 543) respectively). No difference in maximum peak height was found for TAA. Eleven individual AAs showed a significantly 21.8 – 33.6% lower maximum peak height for the gel treatment compared to the unheated drink.

In addition, the time to maximum peak for TAA absorption was 18.8 and 18.9 minutes later for the gel treatment compared to the unheated and heated drink respectively (66.9 min (CI: 59.2 – 74.6) compared to 48.0 min (CI: 37.9 – 58.2) and 48.0 ± 8.4 min (CI: 41.0 – 55.0) respectively). For EAA, time to maximum peak was significantly delayed by 25.4 and 28.8 minutes for the gel treatment compared to the unheated and heated drink respectively (75.4 min (CI: 66.8 – 83.9) compared to 50.0 min (CI: 39.9 – 60.0) and 46.6 min (CI: 38.4 – 54.9)) (**Supplementary Figure 5**). Of the 19 individual AAs measured, 14 AAs showed a 14.4 – 30.3-minute later time to maximum peak height for the gel treatment compared to both drinks.

### 3.2. Gastric emptying

Baseline gastric volume was 36.2 ± 19.5 mL for the gel treatment, 26.3 ± 26.7 mL for the unheated and 40.0 ± 26.4 mL for the heated drink (p = 0.373). An example time series for the unheated drink and the gel treatment is shown in **Figure 4**.

**Figure 4.**
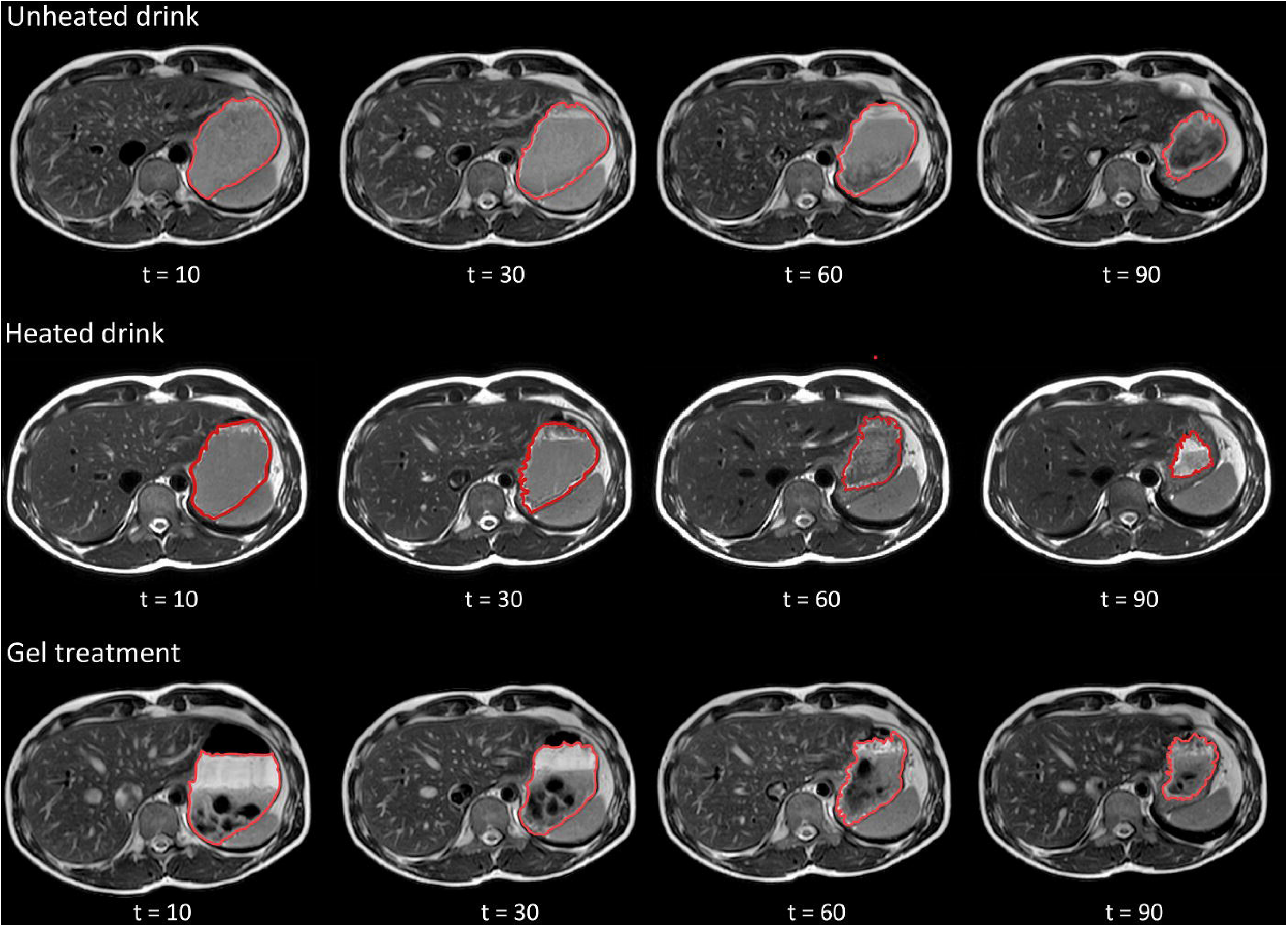
Illustration of gastric emptying over time showing MRI images for the unheated drink and the gel treatment. The stomach content is delineated in red.

**Figure 5** shows an almost linear emptying for the drinks, while the gel treatment shows a quick initial emptying. There was a significant treatment effect on gastric volume over time for the gel treatment compared to the drinks, with a lower volume for the gel treatment (p = 0.002). This effect was driven by timepoints t = 15 up to 70 minutes.

**Figure 5.**
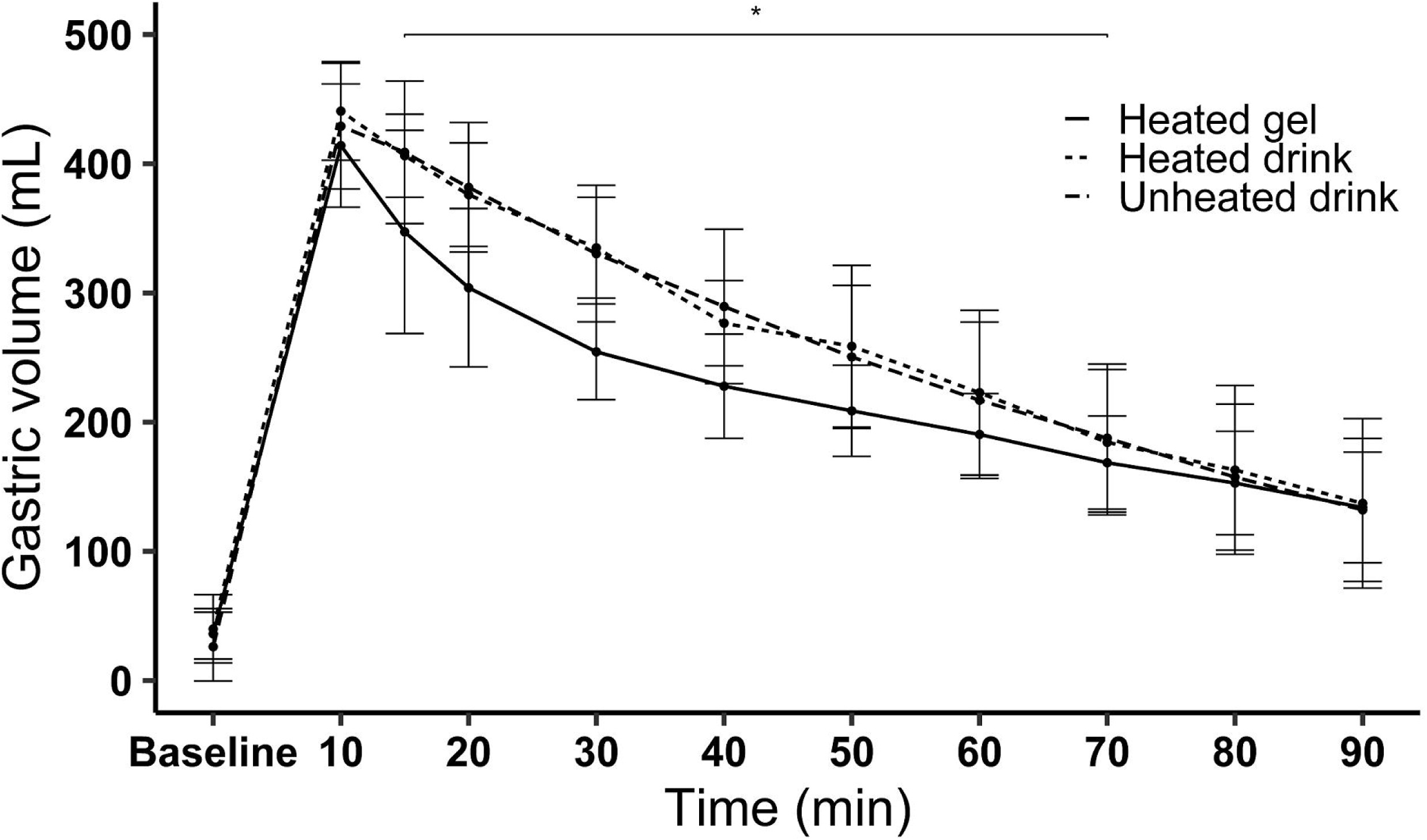
Mean ± SD gastric volume over time of pea protein products. *p < 0.05, as analyzed with a linear mixed model and Tukey HSD correction for multiple comparison. There was a significant treatment effect for the gel treatment compared to both drinks at t = 10 until t = 70 min.

AUC of gastric volume over time showed a trend toward a treatment effect (p = 0.071). On average, AUC of the gel treatment was 16% and 15% lower compared to the heated and unheated drink respectively (17608 ± 3059 mL*min compared to 21037 ± 3999 mL*min and 20605 ± 3892 mL*min, p = 0.086 and 0.149, respectively) (**Figure 6**). There was no difference between the two drinks (p = 0.959).

**Figure 6.**
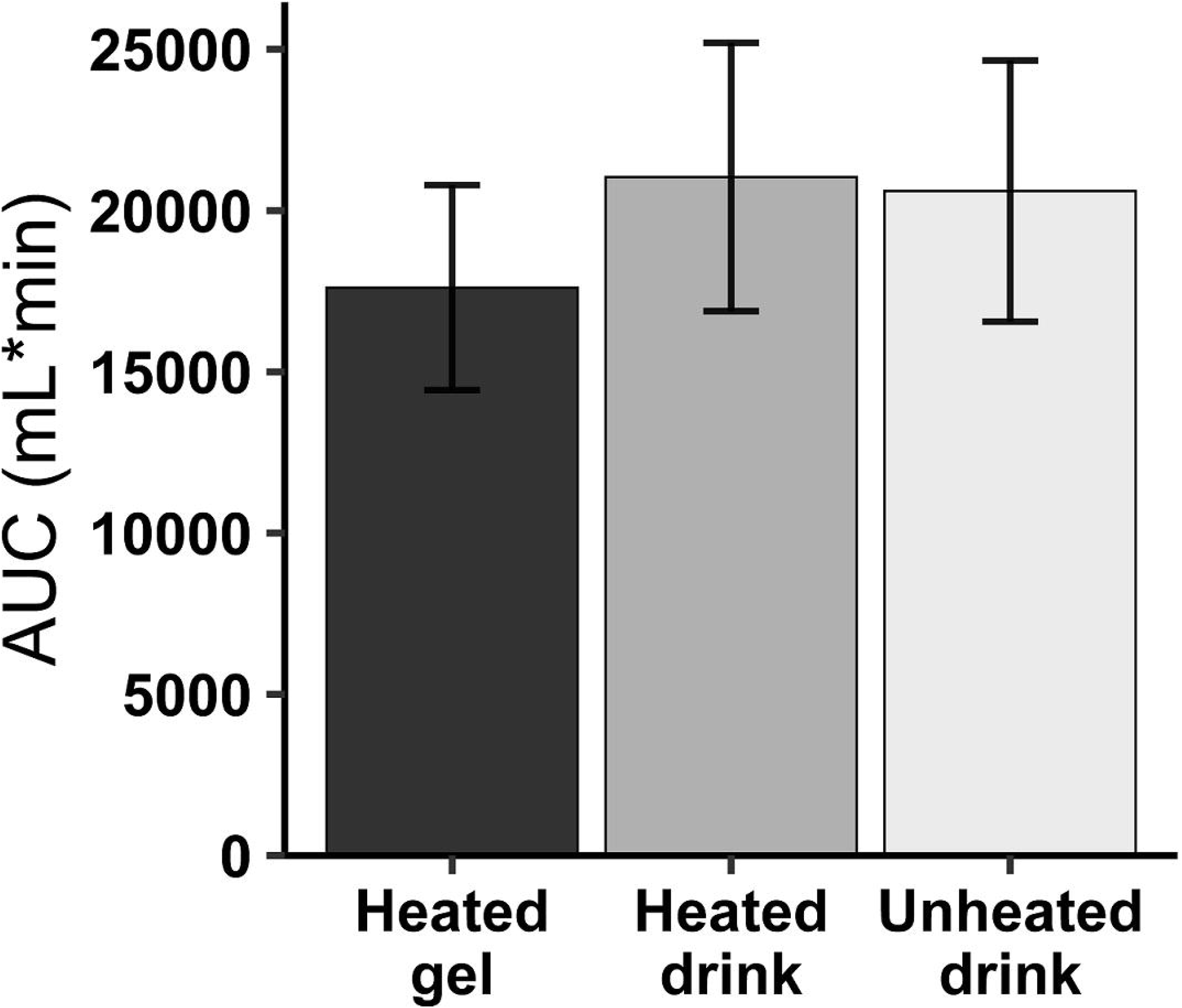
Average AUC ± SD of gastric volume over time for the three treatments. One-way ANOVA showed no significant difference between treatments.

**Figure 7** shows that for the gel treatment the liquid content of the stomach emptied quickly during the first 30 minutes, while the solid content emptied slower. Over 90 minutes, the liquid volume decreased from 300 ± 15 mL to 66 ± 8 mL (78.1% decrease). The solid volume, that is, the protein gel fraction, decreased from 103 ± 5 mL to 69 ± 9 mL (32.4% decrease). For the unheated and heated drink the decrease of liquid volume over 90 minutes was 69.2% and 68.9% respectively.

**Figure 7.**
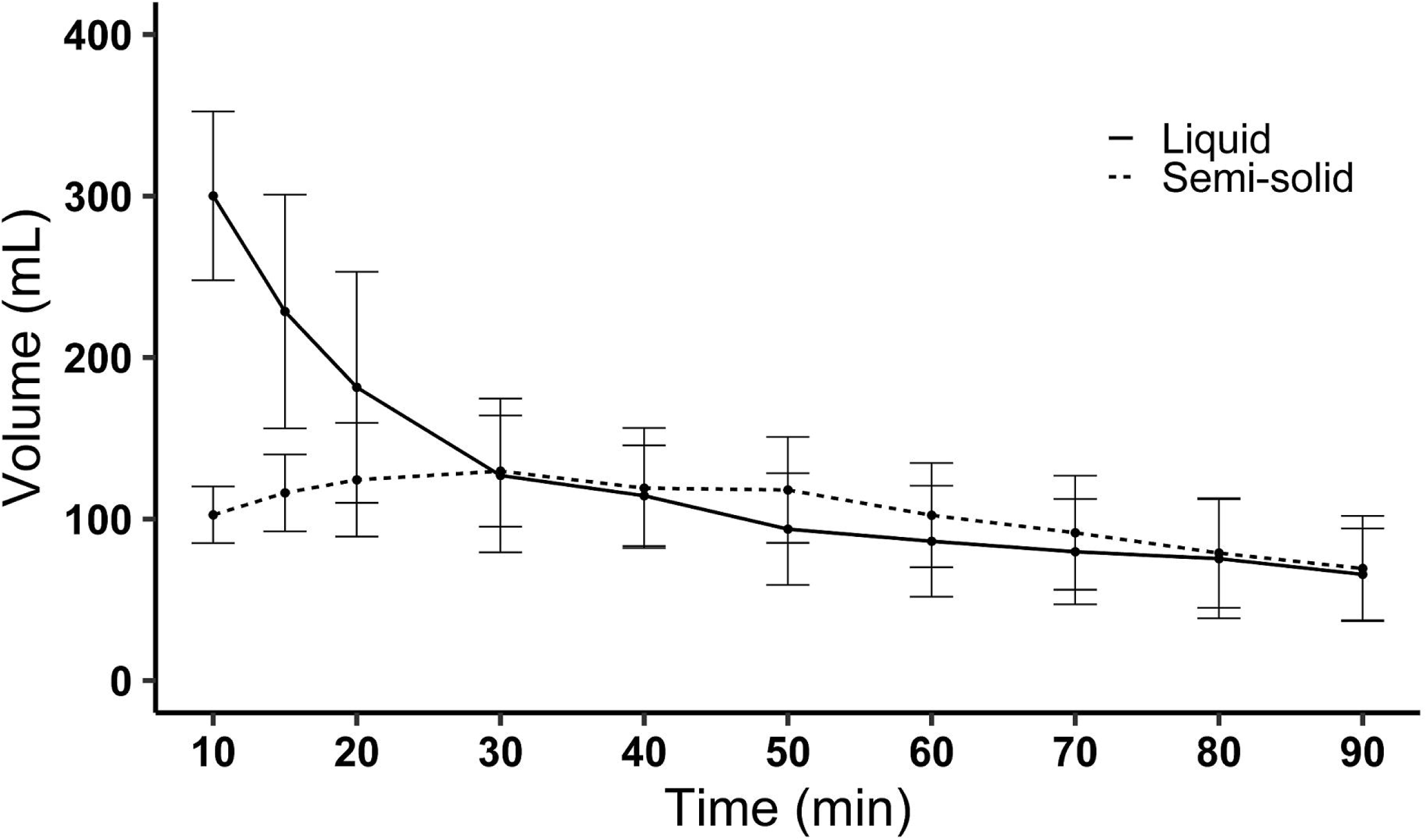
Mean ± SD liquid and semi-solid gastric volume over time of the gel treatment after ingestion of 105 g of pea protein gel with 315 mL water.

### 3.3. Gastric behavior

**Figure 8** shows an overall change in the texture metrics over time (all p < 0.001), where busyness and homogeneity decreased over time and coarseness and contrast increased over time for both drinks. This suggests an increase in the degree of coagulation.

**Figure 8.**
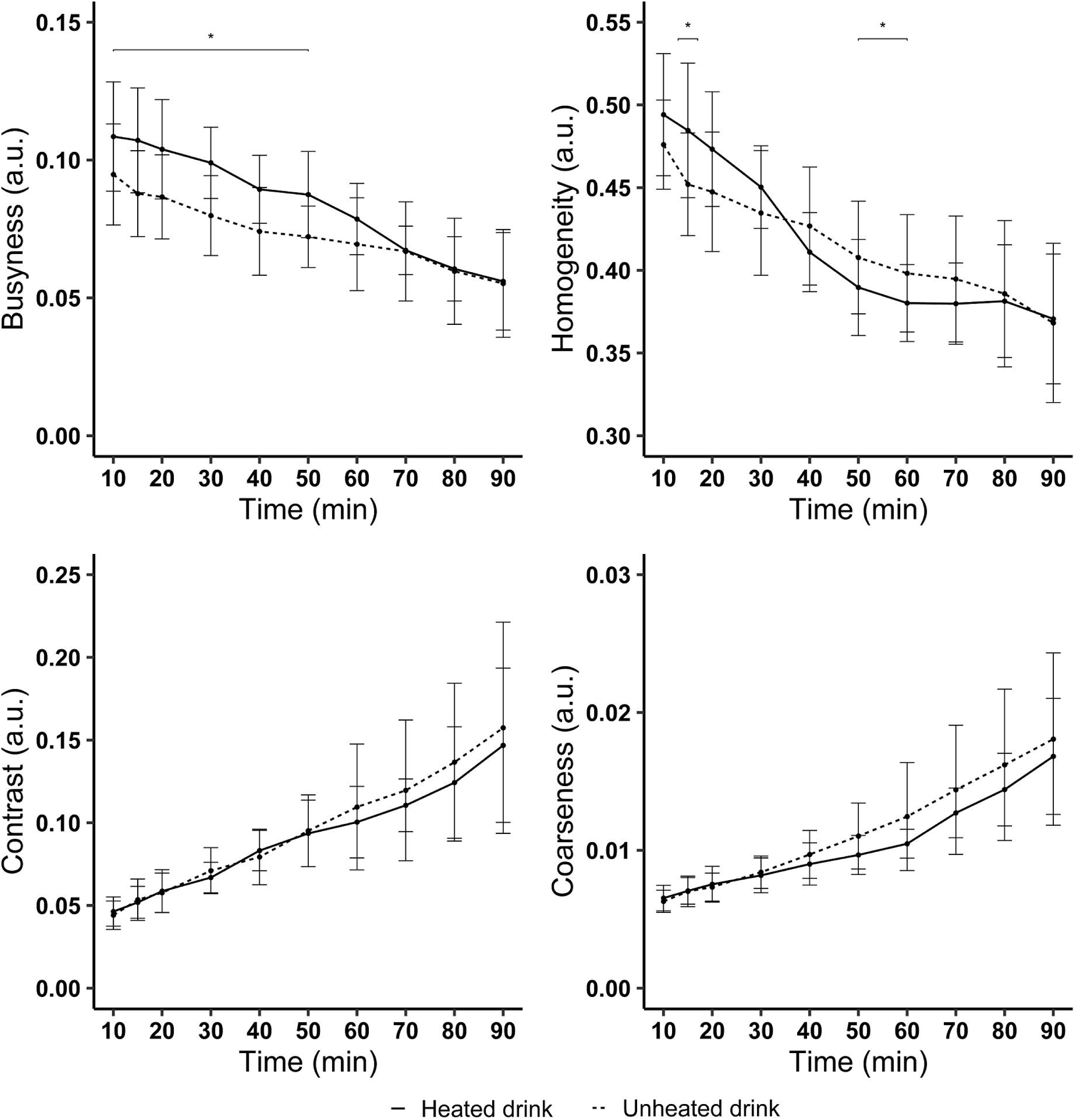
Mean ± SD texture metrics (arbitrary units) of the stomach contents over time for the unheated and heated drinks. *P < 0.05, as analyzed with a linear mixed model and Tukey HSD correction for multiple testing.

No treatment or treatment by time interaction effect was found for contrast (p = 0.204 and p = 0.973) and coarseness (p = 0.295 and p = 0.564). However, for homogeneity a treatment by time interaction was found (p = 0.002). It was lower for the unheated drink at t = 15 min, while it was higher at t = 50 and 60 min compared to the heated drink. Treatment by time interaction was also significant for, with higher values for the heated compared to the unheated drink (p = 0.019). This was driven by timepoints t = 10 until t = 50 min.

### 3.4. Glucose and Insulin

For glucose concentrations there was a trend towards lower concentrations for the unheated drink (p = 0.069). However, the interaction with time was not significant (p = 0.602) (**Supplementary Figure 8**). There was a trend towards lower insulin concentrations over time for the gel treatment (p = 0.058), driven by t = 30 min (**Supplementary Figure 9**).

### 3.5. Appetite and nausea

The graphs of the appetite and nausea ratings are shown in **Supplementary Figure 10**. There was a treatment effect for hunger, fullness, desire to eat, prospective consumption (p < 0.001, p = 0.018, p = 0.003 and p < 0.001, respectively). Hunger (MD -8.3 and -7.1 respectively, p < 0.001), desire to eat (MD -6.7 and -5.2 respectively, p < 0.001) and prospective consumption (MD -7.3 and -8.3 respectively, p < 0.001) were all lower for the gel treatment compared to the unheated and heated drink. Fullness was higher for the gel treatment compared to the unheated drink (MD 4.8, p = 0.014), but not the heated drink (MD 3.5, p = 0.108). However, interaction with time was not significant for hunger, fullness, desire to eat and prospective consumption (p = 0.714, p = 0.960, p = 0.999 and p = 0.998, respectively). Thirst did not differ between treatments (treatment effect: p = 0.359, treatment by time interaction p = 0.998). Nausea showed a treatment effect (p = 0.003) with lower levels for the gel treatment compared to the unheated drink (MD -2.9, p = 0.002), but not the heated drink (MD -1.5, p = 0.230). However, there was no interaction with time (p = 0.283).

### 3.6. *In vitro* static digestion

**Figure 9** shows the *in vitro* degree of hydrolysis of the three treatments for 2 hours of gastric digestion (0 – 120 min) and 2 hours of intestinal digestion (120 – 240 min). The digestibility, expressed as degree of protein hydrolysis of the unheated drink was slightly higher compared to that of the heated drink and gel treatment during the gastric phase (9.6% compared to 5.2% and 3.0% at 120 min, respectively). In the intestinal phase, the gel treatment had a higher degree of hydrolysis compared to the unheated and heated drinks (57.6 compared to 38.3 and 38.9% at 240 min respectively). Precipitation of the drinks was similar at t = 0. However, for the heated drink precipitation increased over time, while it was stable for the unheated drink (**Supplementary Figure 11**). This was confirmed with the semi-dynamic digestion model (**Supplementary Figure 12**). Moreover, the heated drink had larger particles compared to the unheated drink (**Supplementary Figure 13**). Supplementary Figure 13-B and 13-C show the volume density of the particles in both drinks at 0, 60 and 120 minutes after the start of digestion. In both drinks, most particles were around 10 µm. Both drinks showed a decrease over time for larger particles, while the volume density for smaller particles increased. The maximum particle size for the unheated drink was 270 µm, while the heated drink showed particles sizes up to 500 µm. The cocoa powder showed a large peak around 24 µm and a small peak around 1 µm.

**Figure 9.**
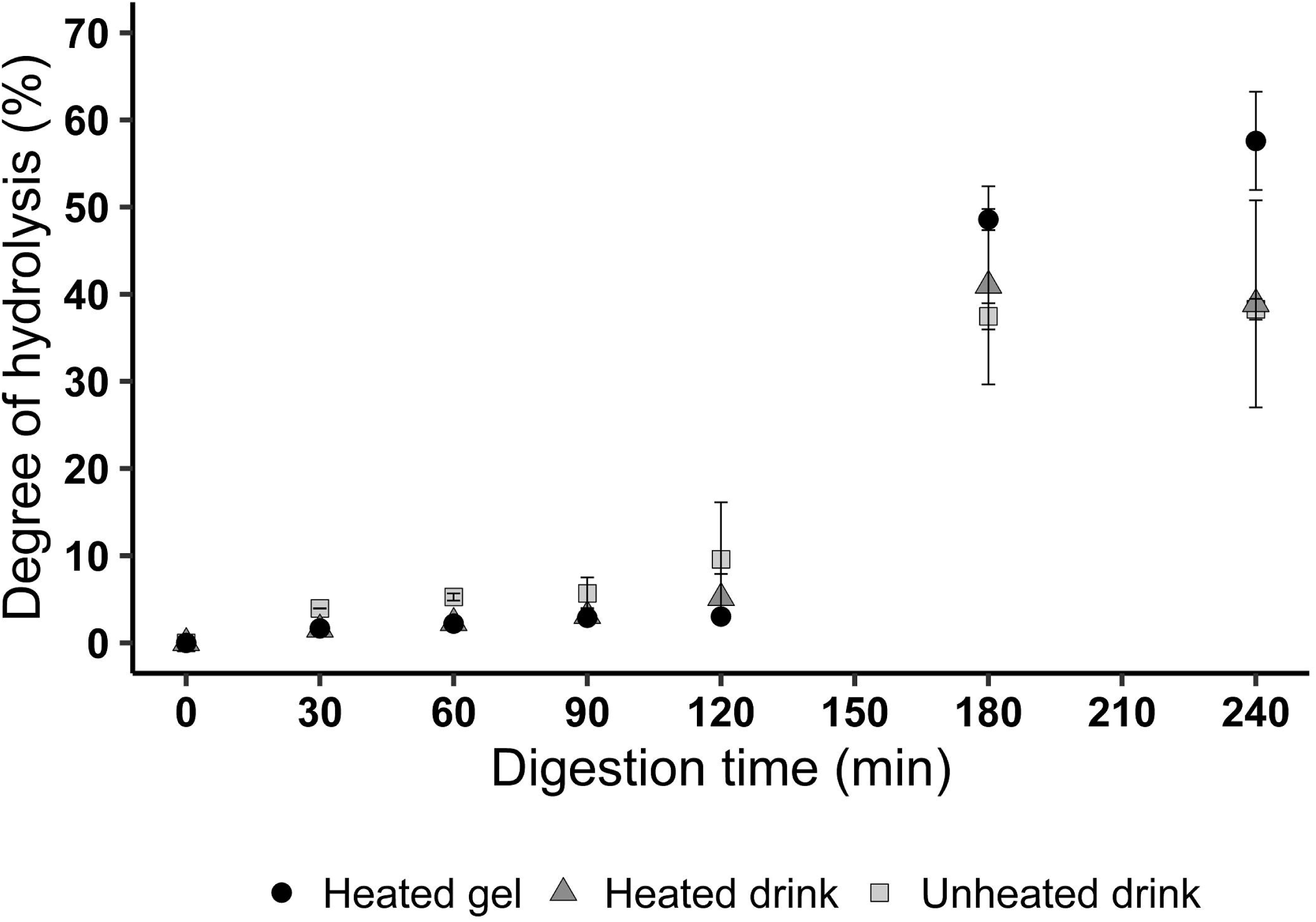
Degree of hydrolysis (%) of the three pea protein products measured via a static in vitro protocol.

Moreover, in the gastric phase (0-120 min), the heated drink showed a higher number of soluble peptides (**Supplementary Figure 14**). In addition, for the gel treatment it took about an hour in the gastric phase until the same amount of dissolved peptides was present. During the intestinal phase (180 and 240 min), the AUC was higher for the gel treatment compared to the unheated and heated drink (8415 compared to 5660 and 5791 mAu*min at 240 min, respectively), which is in agreement with the higher degree of hydrolysis (Figure 9). For the drinks, the number of large molecules decreased over digestion time and more small size peptides became soluble (**Supplementary Figure 14**).

## 4. Discussion

This study assessed gastric behavior and subsequent AA kinetics for three iso-caloric and iso-volumetric pea protein products differing in heat treatment and texture. Heat treatment did not affect gastric emptying, but texture did. A gel consumed together with water showed overall faster initial emptying compared to a drink. This initial quick emptying was attributed to passage of the water, while the gel fraction emptied more slowly. Consequently, the time to maximum peak for AA was delayed by 18.8-28.8 minutes for the gel treatment compared to the two drinks.

The industrial processing required to manufacture pea protein isolate includes heat treatment. Since the heat-treated drink did not show altered gastric emptying or AA absorption kinetics, this confirms that additional heat treatment did not further affect digestibility. *In vitro* work by Rivera del Rio, et al. (2020) showed that heat treatment of pea protein isolate not only results in small and suspended particles that can be better hydrolyzed by pepsin in the stomach but also induces aggregates, which are less digestible. Thus, although they found that heat treatment of pea protein isolate affects the structure of the proteins, it did not significantly affect the overall in vitro gastric digestibility, which is in line with our findings.

Gastric volume was lower for the gel treatment in the first hour after consumption compared to the drinks. Consumption of a semi-solid gel with water resulted in an initial quick emptying of the watery contents and slower emptying of the gel compared to the drinks (32% of the gel had emptied at t = 90 min, compared to 69% of the drinks). This is in line with previous research of Mackie, et al. (2013) who found slower gastric emptying after consumption of a semi-solid compared to an iso-caloric liquid meal containing animal-based proteins (grated gouda cheese and low-fat yogurt consumed with water compared to a homogenous liquid mixture of sunflower oil, sodium caseinate, whey protein isolate and sugar). In addition, Marciani, et al. (2012) showed that when the solid and water fraction are not homogenized, the water sieves past the gastric content and empties quickly. When the same meal was blended into a soup, gastric content volume decreased more slowly in a linear fashion (Marciani, et al., 2012). This is in line with our results for the pea protein drinks, which had an approximately linear emptying curve. In addition, the lower accessibility of pepsin to penetrate a food bolus explains why hydrolysis of a semi-solid protein food was slower compared to that of the protein drinks, leading to slower gastric emptying (Bornhorst, et al., 2016; Luo, Boom, & Janssen, 2015). No clear correlation was found between the slower emptying of the pea protein gel and AA absorption kinetics. However, the delay that was found in gastric emptying was also reflected in the absorption of AA in the blood, which showed an 18.8 – 28.8 min delay and lower maximum peak compared to the drinks. This is in line with our expectation that delayed gastric emptying results in delayed AA absorption.

The attenuated rise in postprandial AA concentrations after consumption of a solid versus liquid form was reported in multiple studies (Conley, et al., 2011; de Hart, et al., 2021; Hermans, et al., 2022; A. M. H. Horstman, et al., 2021). However, these studies compared products that did not only differ in texture, but also macro- and micronutrient composition, protein composition and/or volume. In contrast, van Lieshout, et al. (2023) compared liquid vs solid iso-caloric and iso-volumetric products based on whey isolate and calcium caseinate and found no difference in postprandial AA concentrations in health females. This difference might be explained by the nature of the proteins. Animal-based proteins, especially caseins, are known to coagulate, thereby delaying gastric emptying (Huppertz & Chia, 2021) and AA absorption kinetics (A. M. Horstman & Huppertz, 2022).

Although no coagulates were visible on the MRI images, all four texture metrics showed a change over time that suggests a higher degree of coagulation over time for both drinks. That is, lower homogeneity and busyness over time and a higher contrast and coarseness. Moreover, based on busyness and homogeneity, one might conclude that the unheated drink showed a greater degree of coagulation in the first 60 minutes compared to the heated drink. This is in contrast to our *in vitro* findings where the unheated drink did not show an increase in precipitation over time, while the heated drink did. In literature, the results of *in vitro* digestion research on pea protein coagulation are also inconsistent. An *in vitro* study by Overduin, Guérin-Deremaux, Wils, and Lambers (2015) showed that a 3% solution of the same pea protein isolate as used in this study forms coagulates of 50-500 µm within 2 hours. This is in agreement with our measurements that showed a maximum particle size of 500 µm. Coagulates of this size will not be visible on the MRI images, with a resolution of 1 by 1 by 4 mm. However, formation of such small coagulates could still affect the intensity of these T_2_-weighted scans. This might explain the observed changes in the image texture metrics. In addition, since these texture metrics look at intensity contrast in the stomach, gastric juice might influence these metrics, since it appears as a high image intensity. This requires further validation. However, even when these small coagula would be present, it is not likely to affect gastric emptying since particles <1-2 mm can be emptied through the pylorus (Kong & Singh, 2008). This is in line with our results, where no differences in gastric emptying were observed between both drinks. In addition, AA absorption kinetics were also not different. Based on these findings we conclude that even if pea protein isolate coagulates in the stomach, this does not affect further digestion.

Our results indicate that consumption of a semi-solid food results in increased feelings of satiety compared to the consumption of iso-caloric and iso-volumetric liquid foods. This is in contrast to a study of Marciani, et al. (2012) that showed that a mixed solid/liquid food is less satiating compared to the same meal in homogenized form (AUC of hunger, 1166 ± 76 compared to 1106 ± 65 VAS score*min (mean ± SEM), P < 0.02). They suggested that the lower satiation might be due to the quick initial emptying of the liquid portion of the meal, which reduces gastric volume and thus lowers sensation of fullness. However, a study of Zijlstra, et al. (2009) comparing liquid and semi-solid texture found that consuming semi-solids was more satiating, even though there was no difference in CCK-8 or GLP-1 responses (fullness P = 0.03, desire to eat P = 0.04). Camps, et al. (2016) also showed that increasing viscosity increased satiation and satiety. One explanation for this is the greater degree of oral exposure when consuming the gel. The mean ingestion time was 1.4 times longer for the gel treatment compared to the drinks. In addition, participants consumed the drink with a straw, while they needed to chew on the gel. Longer mastication for an isocaloric load leads to higher feelings of satiety (Forde & Stieger, 2022; Lasschuijt, de Graaf, & Mars, 2021; Wanders, et al., 2013). However, it should be noted that overall differences between the semi-solid treatment and drinks were small, with a mean difference <10, which is often considered as a cut-off point for clinical relevance (Flint, Raben, Blundell, & Astrup, 2000).

This study used MRI to examine gastric behavior. This requires participants to be scanned in a supine position. Although the effect is small, studies have shown that protein ingestion in an upright sitting position accelerates gastric emptying and increases the postprandial rise in plasma AA availability by increasing protein digestion and AA absorption rates compared to a supine position (Holwerda, Lenaerts, Bierau, & Van Loon, 2016; Holwerda, Lenaerts, Bierau, Wodzig, & van Loon, 2017; Jones, et al., 2006; Spiegel, et al., 2000). The study of Holwerda, et al. (2017) showed a higher peak plasma leucine concentration for upright sitting compared to a supine position (213 ± 15 compared to 193 ± 12 μmol/L, P < 0.05). However, the participants were scanned in the same position for all treatments. Therefore, the relative differences between treatments are expected to remain the same.

In the *in vitro* digestion the unheated drink showed a slightly higher degree of hydrolysis in the gastric phase, while the gel treatment showed an increased degree of hydrolysis during the intestinal phase. This suggests, that in the stomach, the gel structure reduces the access of pepsin. However, after two hours in the gastric phase the gel might have swollen, leading to a looser structure that is more accessible for trypsin in the intestine. In addition, the peristaltic movements in the intestine might result in increased fractionation, allowing for a larger surface area. This is in agreement with the *in vivo* results, where the gel treatment showed lower AA concentrations during the first ∼60 minutes, but comparable concentrations after that.

The heated drink showed higher concentrations of soluble peptides at the start of digestion. The higher solubility of the heated drink is likely a result of the heating process (Rivera del Rio, Möller, Boom, & Janssen, 2022). In addition, both drinks showed precipitation at the start, which can be explained by the low solubility that plant-based proteins are known for. However, in both the static and semi-dynamic digestion the heated drink visually showed more sedimentation over time, indicating higher levels of aggregation. This was also confirmed with the particle size distribution measurements, which showed that the heated drink had larger particles.

To conclude, this study demonstrates that heat treatment of pea protein isolate does not affect gastric emptying or AA absorption. However, consuming pea protein isolate in a product with a semi-solid texture slowed down both gastric emptying and subsequent AA absorption compared to liquids, but did not affect total absorption kinetics. These results suggest that texture influences the rate at which pea protein is absorbed, but not total absorption. In addition, comparison with *in vitro* data showed that *in vitro* digestion models gave additional support and insight to *in vivo* digestion results.

## Data Availability

All data produced in the present study are available upon reasonable request to the authors

## List of abbreviations

AA: Amino Acid
DIAAS: Digestible Indispensable Amino Acid Score
EAA: Essential Amino Acids
SGF: Simulated Gastric Fluid
TAA: Total Amino Acids

## Acknowledgements and statement of authors’ contributions to manuscript

PS, AJ, EE and DE: conceptualization, methodology; JR, PP, JJ: investigation; JR, JJ and RW: data curation; JR and RW formal analysis; JR Writing - Original Draft; EE, PP, JJ, DE, AJ and PS: Writing - Review & Editing; JR: visualization; PS had primary responsibility for final content. All authors read and approved the final manuscript. None of the authors declare a conflict of interest. We thank Caya Lindner for assisting with data collection. The use of the 3T MRI was made possible by WUR Shared Research Facilities.

