## Supplement for "Gastric emptying and nutrient absorption of pea protein products differing in heat treatment and texture: a randomized *in vivo* crossover trial and *in vitro* digestion study"

**Methods – Product preparation**

Pea protein gel

Ingredients (104.5g):

- 25 g pea protein isolate (Nutralys® F85M)
- 60 mL water
- 8 pcs cyclamate and saccharin sweeteners (Kruger)
- 7 mL vanilla aroma (Dr. Oetker)
- 5 mL chocolate aroma (Nielsen Massey Chocolate-extract)
- 7.5 g cacao

Preparation:

- Combine and thoroughly mix all ingredients
- Cover with plastic foil and poke a few holes in the plastic
- Steam it in the steam oven for 30 min on 90 °C

Pea protein drink

Ingredients (419.5g):

- 25 g pea protein isolate (Nutralys® F85M)
- 60 mL water
- 8 pcs cyclamate and saccharin sweeteners (Kruger)
- 7 mL vanilla aroma (Dr. Oetker)
- 5 mL chocolate aroma (Nielsen Massey Chocolate-extract)
- 7.5 g cacao

Preparation:

- Combine and thoroughly mix all ingredients

Heated drink:

- Cover with plastic foil and poke a few holes in the plastic
- Steam it in the steam oven for 30 min at 90 °C

**Supplement**

**Methods – In-vitro digestion model**

The three treatments were analyzed using the static *in vitro* digestion model. For the drinks, a load of 5 mL was used and for the gel treatment 1.25 g of gel was used together with 3.75 mL water.

*Gastric phase*

The volume ratio of drink to SGF (simulated gastric fluid) included pepsin (2000 U/mL final activity) was 1:1 for the gastric phase of the digestion. The gastric digestion was performed for 2 hours at 37˚C and separate tubes were used for each sampling time point. After sampling, digestion was stopped by adding a predefined amount of NaOH to reach pH 7 to inactivate pepsin and the tube was placed in an ice bucket.

*Intestinal phase*

This step is a follow-up of the gastric digestion. The gastric digestion was stopped by adding a predefined amount of NaOH to get pH 7.0. Subsequently, SIF (simulated intestinal fluid) was added in a volume ratio of 1:1. Next to that, pancreatin (final trypsin activity 100 U/mL) was added to start the intestinal digestion. The intestinal digestion was performed at 37°C for 2 hours with individual tubes for 60 and 120 minutes. To stop the digestion, the tubes were heated at 95°C for 5 minutes and afterwards placed in an ice bucket.

*Peptide characterization*

High Performance Size-Exclusion Chromatography (HPSEC) was used to analyze the peptide size distribution during digestion. An UltiMate 3000 chromatographic system

(ThermoFischer Scientific Inc., USA) is used with a two TSK gel columns G3000SWXL and G2000SWXL. The eluent consisted of 30% v/v acetonitrile and 0.1% v/v trifluoro acetic acid and the flow rate was 1.5 mL/min. The UV-detector was set at 214 nm. Details on the analysis were described by Rivera del Rio, et al. (2020).

*Degree of hydrolysis*

The degree of hydrolysis is the number of cleaved peptide bonds divided by the total number of peptide bonds, expressed as percentage. It can be calculated using this equation: $DH = \frac{h}{h_{tot}} \cdot 100\%$. Where $h_{tot}$ is the total number of peptide bonds in 1 kg protein in meq/g. The number of peptide bonds cleaved in 1 kg protein, h, can be calculated as follows: $h = \frac{\frac{[NH_{2,free}]}{[Protein]}}{\alpha}$. Where [$NH_2$, free] is the calculated concentration of free amino groups in the samples by using a standard curve, [Protein] is the initial protein concentration and α stands for the relationship between [$NH_2$, free] and the color intensity measured and is 1 for pea protein.

The concentration of free amino groups is measured via a colorimetric reaction with o-phthaldialdehyde (OPA). The method has been described by Rivera del Rio, et al. (2022). The calibration curve was prepared with a leucine standard solution. The experiments were done in duplicate.

*Semi-dynamic in vitro digestion model*

The unheated and heated drink were tested in a semi-dynamic system, where 1.21 mL of SGF (simulated gastric fluid) containing 4000 U/mL pepsin was added at 4 time points: t = 0, 30, 60, 90 min. After adding SGF, the tube was shaken. No gastric emptying was included.

### Supplementary Figures

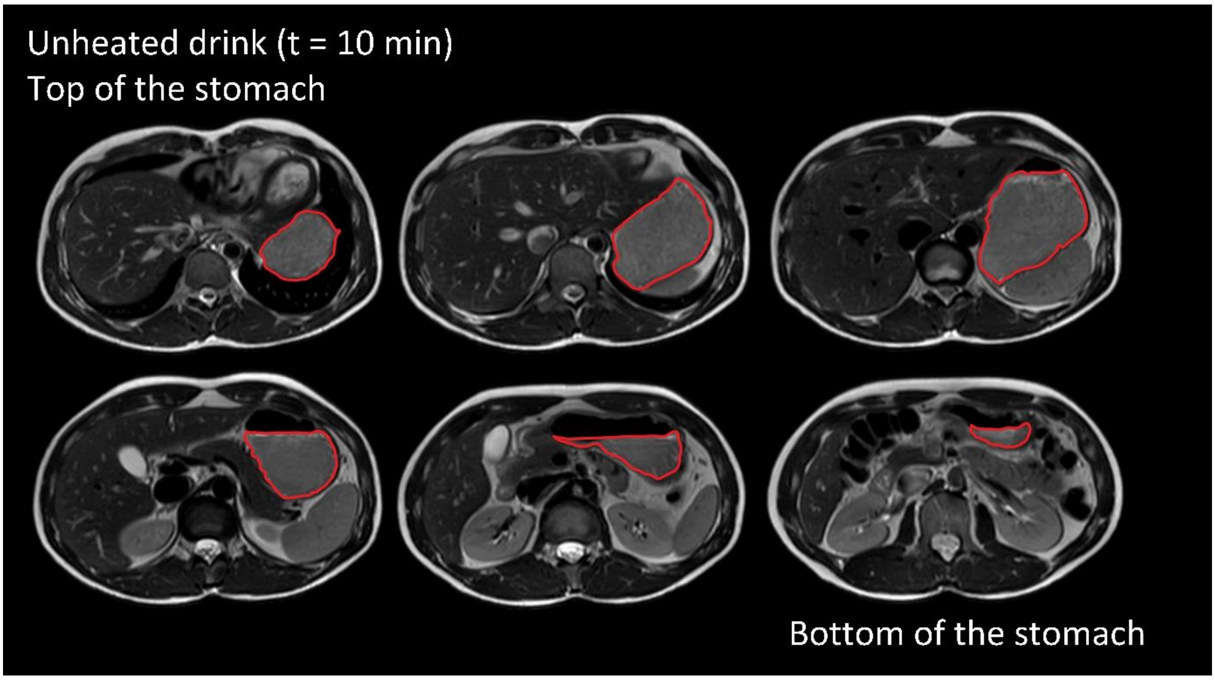


Supplementary Figure 1. MRI scan with delineated stomach after ingestion of the unheated drink (420 mL) at 10 min after the start of ingestion from top to bottom.

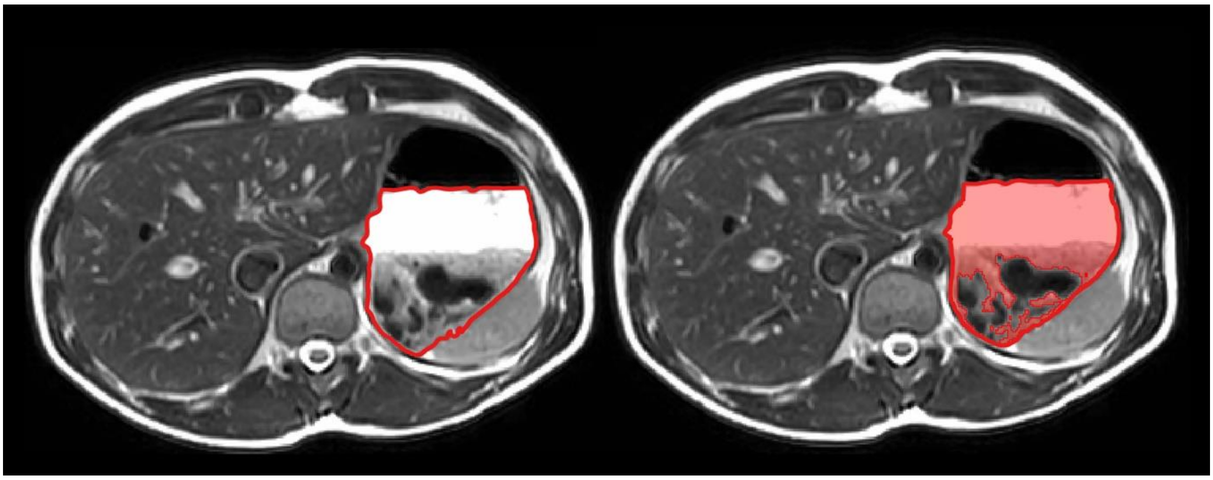

Supplementary Figure 2. Example of two identical $T_2$-weighted MRI images with total gastric volume delineated (left) and total gastric volume with liquid and semi-solid content marked in red (right).

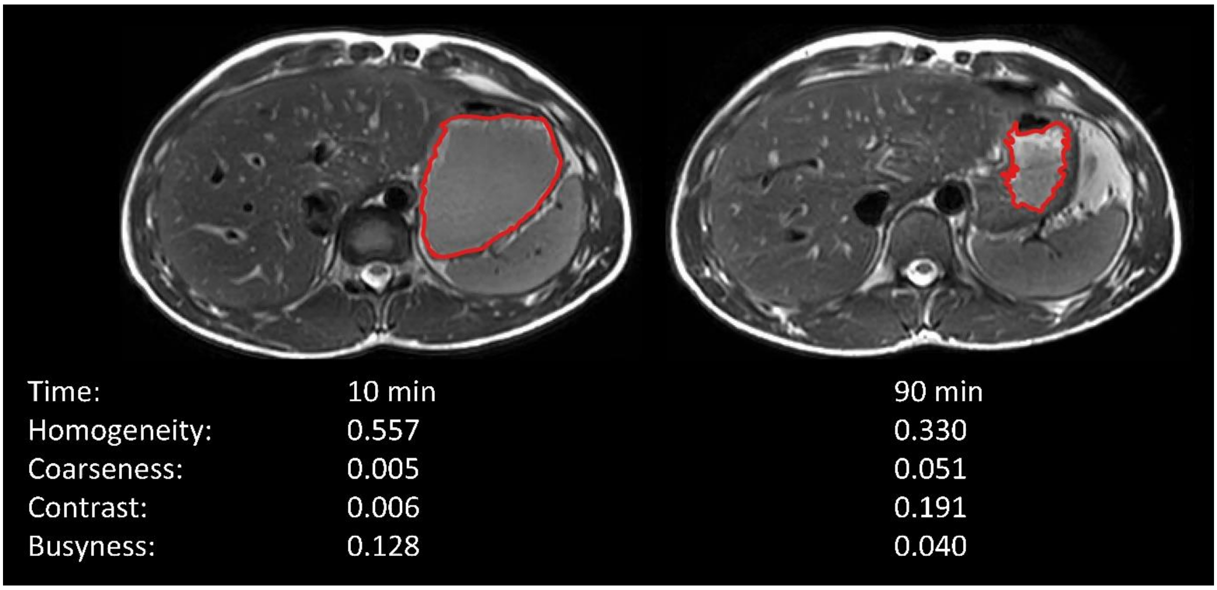


Supplementary Figure 3. Example of two $T_2$-weighted MRI images with their associated image texture metrics indicating relatively low (t = 10 min) and high (t = 10 min) coagulation.

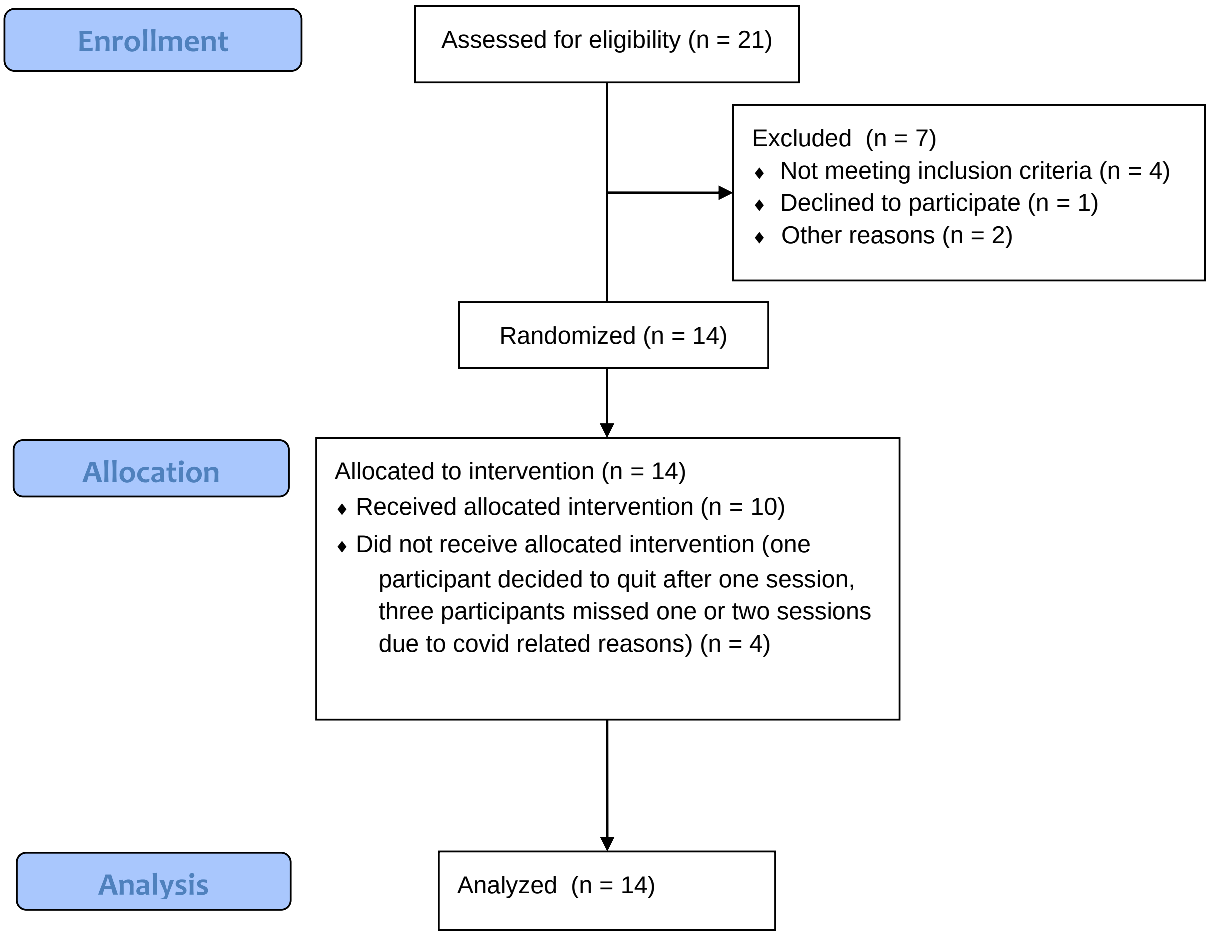


Supplementary Figure 4. Flow diagram for inclusion, treatment allocation and analysis.

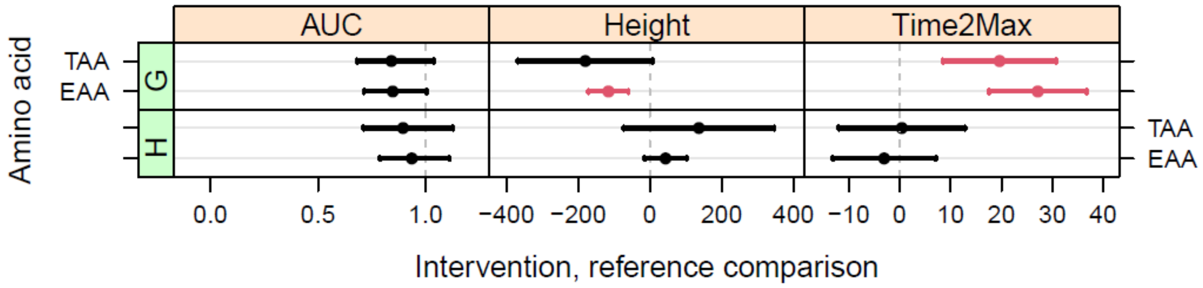


Supplementary Figure 5. Mean differences ± SD in AUC (fold change), maximum peak height (µM) and time to maximum peak (min) of AA of the gel treatment and heated drink compared to the unheated drink. Significant differences are indicated in red. G = gel treatment, H = heated drink, TAA = total amino acids, EAA = essential amino acids.

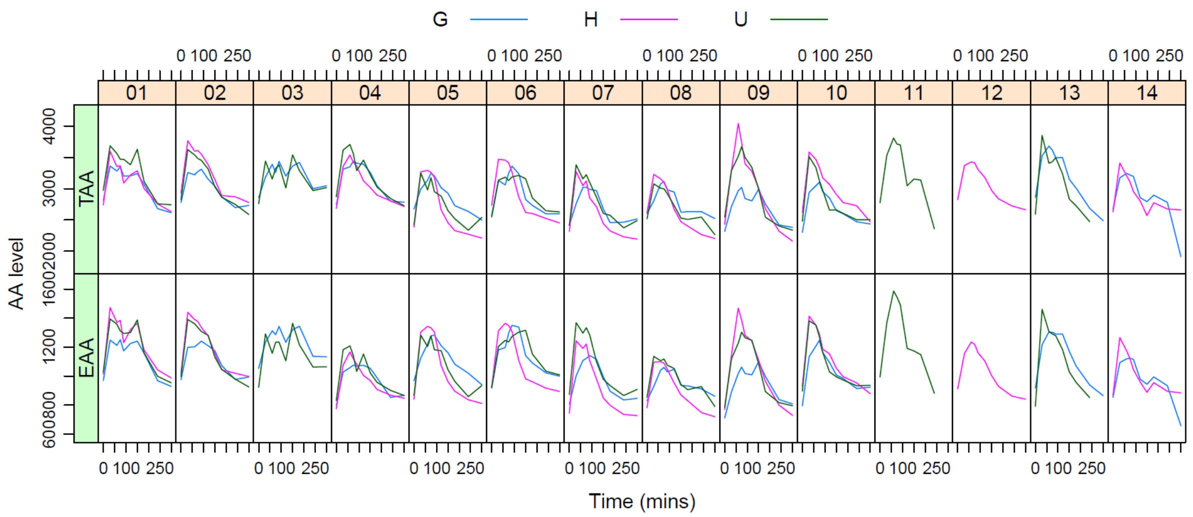


Supplementary Figure 6. Total amino acid (top) and essential amino acid (bottom) levels over time of the participants (01-14) after consumption of the three pea protein products. G = gel treatment, H = heated drink, TAA = total amino acids, EAA = essential amino acids

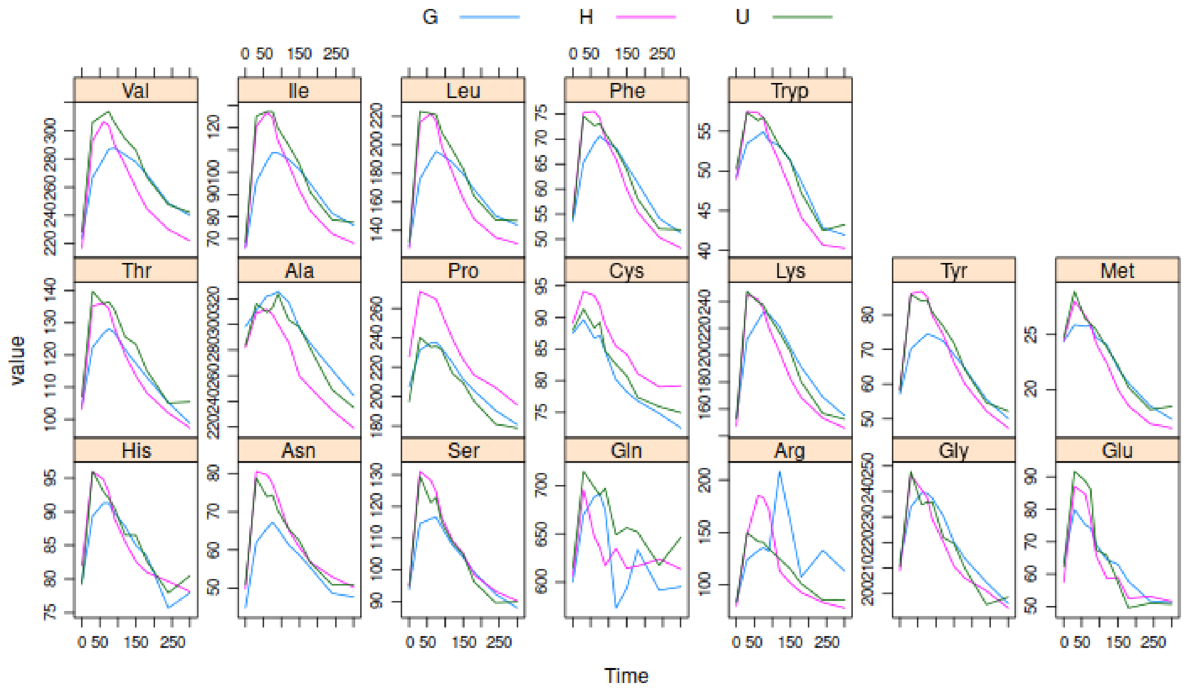


Supplementary Figure 7. Mean amino acid levels over time after consumption of the three pea protein products. G = gel treatment, H = heated drink

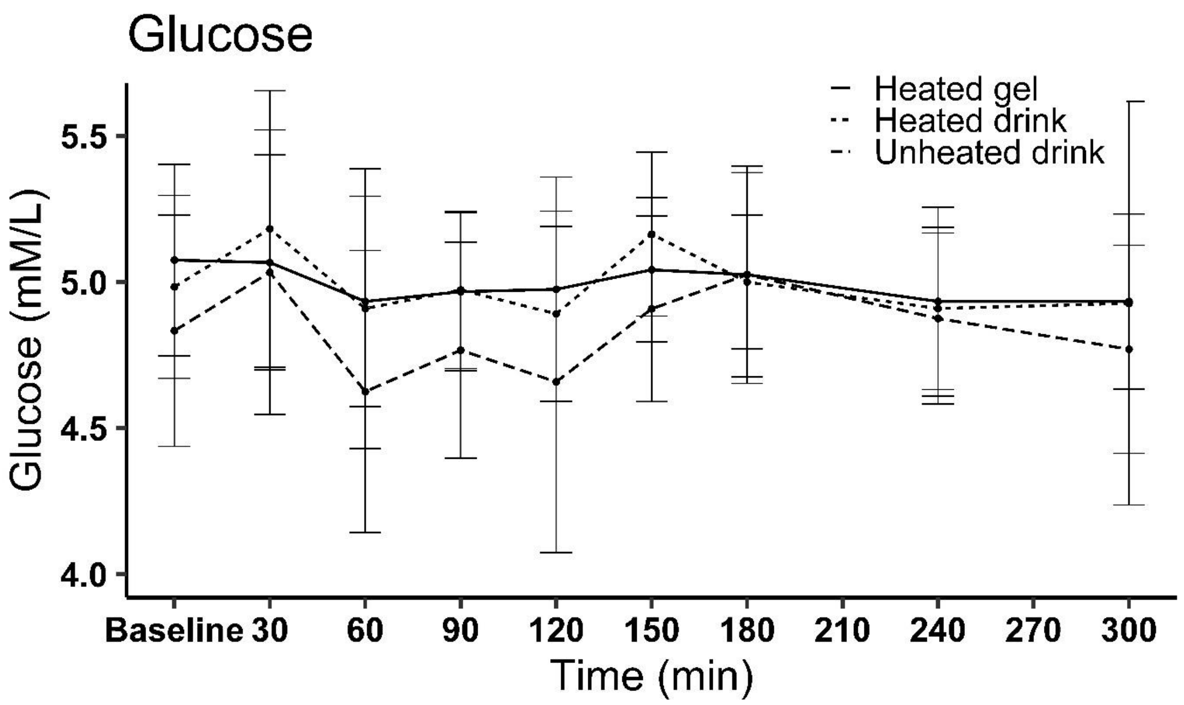


Supplementary Figure 8. Mean ± SD glucose concentration over time for three treatments.

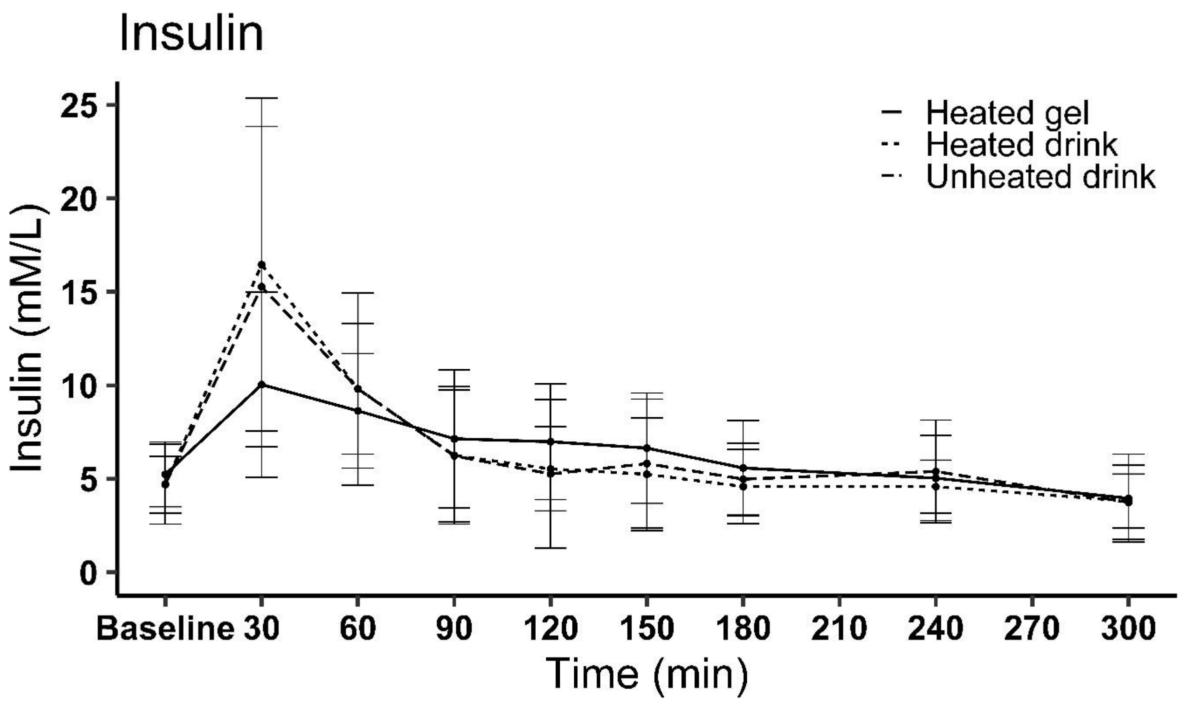


Supplementary Figure 9. Mean ± SD insulin concentration over time for three treatments.

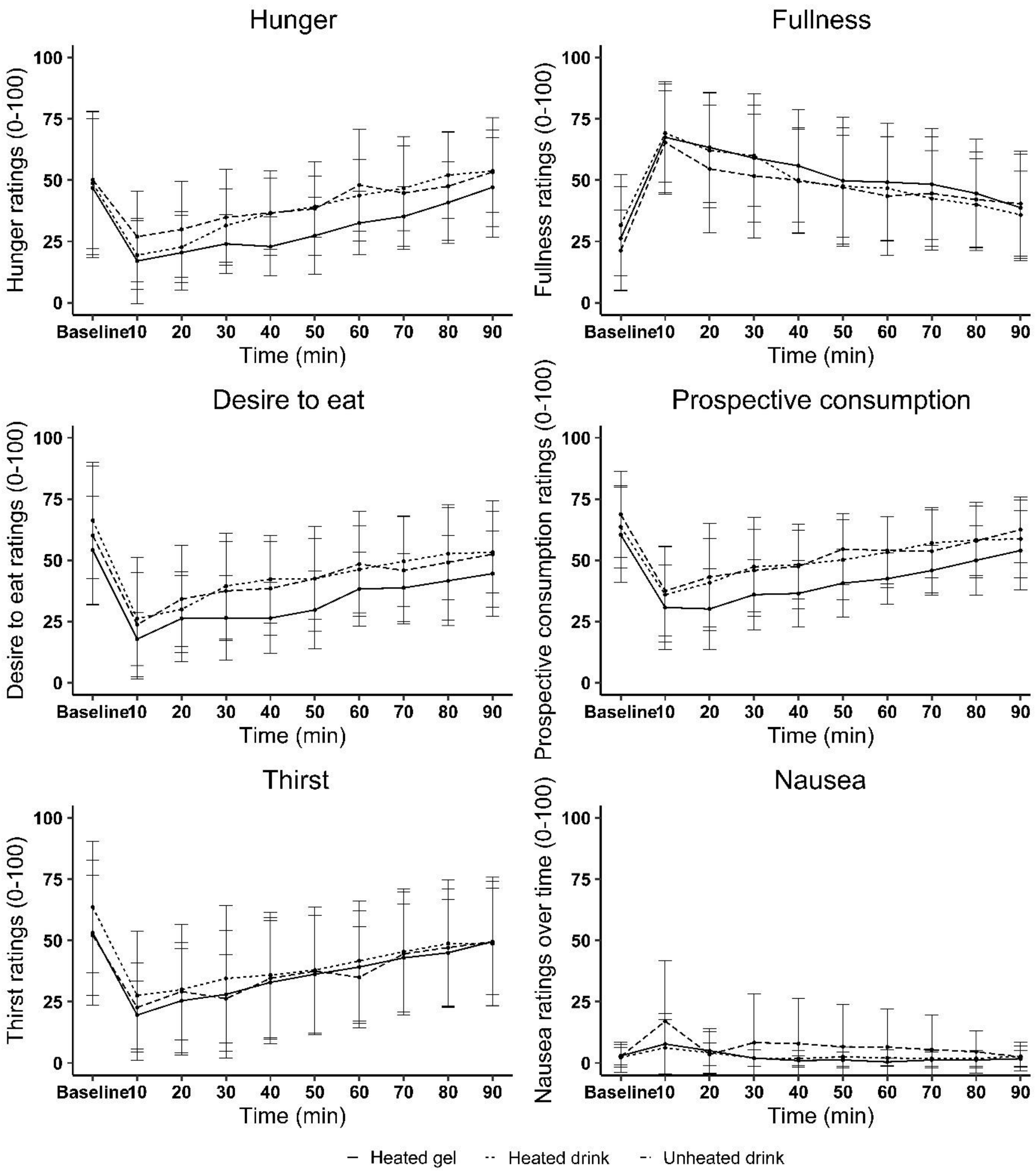


Supplementary Figure 10. Appetite and nausea ratings over time during the test session per treatment. Graphs depicting mean ± SD for hunger, fullness, appetite, expected prospective consumption, thirst and nausea over time after ingestion of pea protein products.

A

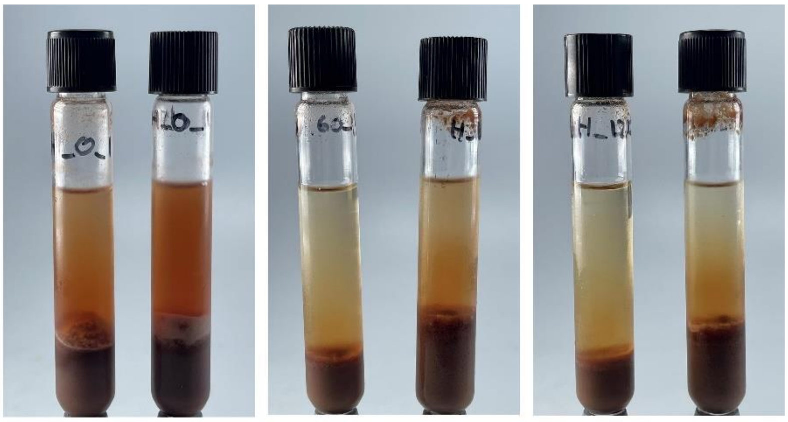

0 minutes 60 minutes 120 minutes

B

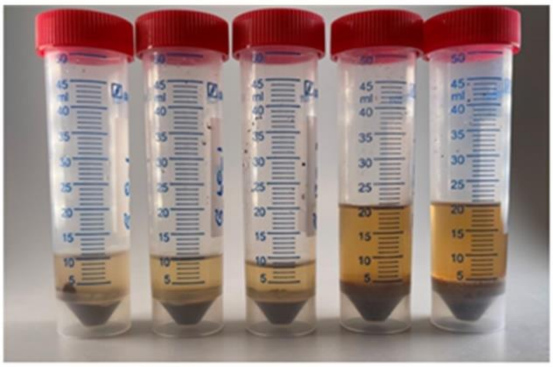

0, 60, 120, 180 and 240 minutes

Supplementary Figure 11. Precipitation during the digestion process: (A) precipitation of the unheated (left tube) and heated (right tube) drinks and (B) precipitation of gel and water 0, 60, 120, 180, 240 minutes from left to right.

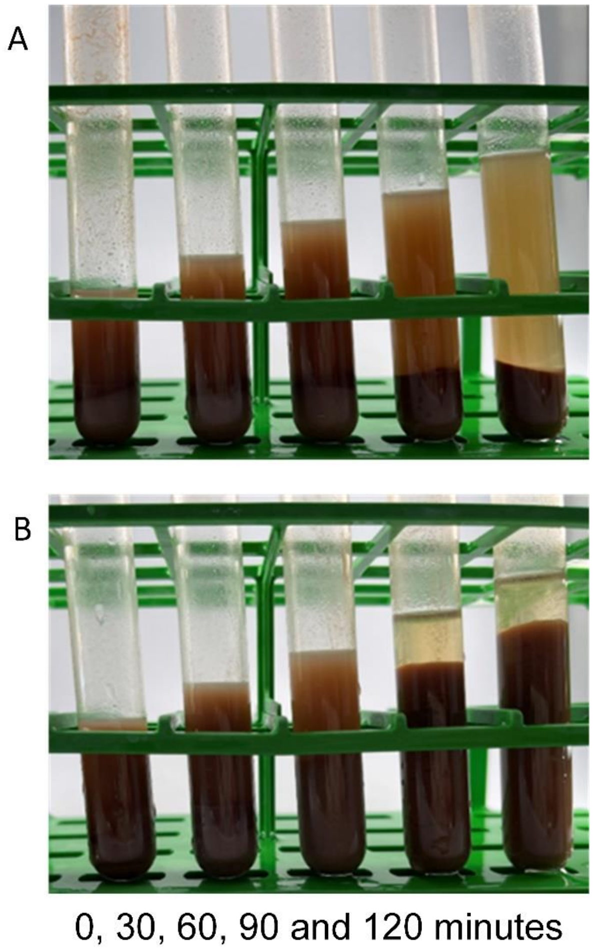


Supplementary Figure 12. Precipitation of unheated (a) and heated (b) drinks over digestion time (0, 30, 60, 90 and 120 minutes from left to right) during semi-dynamic *in vitro* digestion.

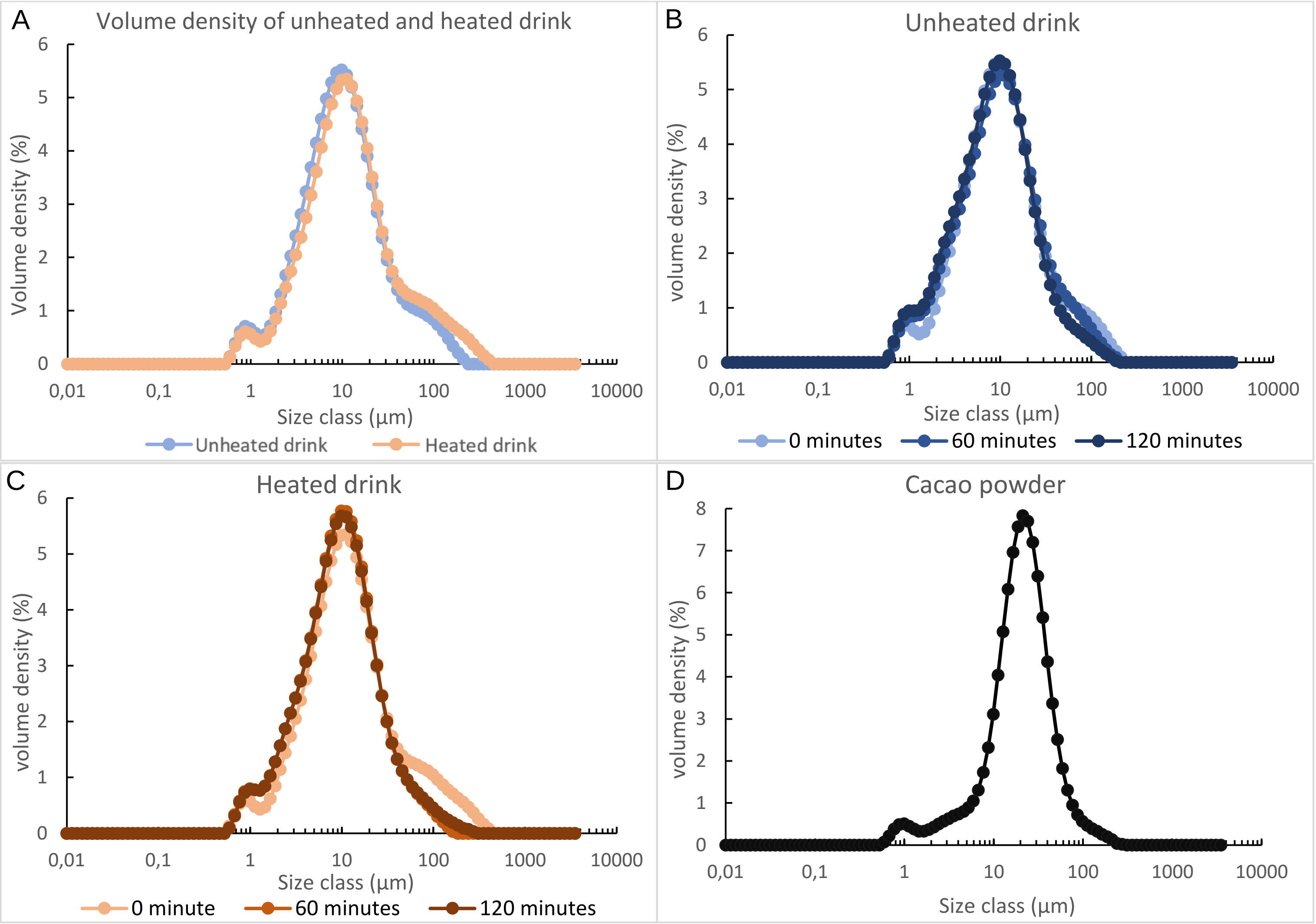


Supplementary Figure 13. Volume density (%) of particle size distribution for the A) unheated and heated drink at the start of digestion (0 min), B) of the unheated drink at 0 min, 60 min and 120 minutes after the start of digestion, C) of the heated drink at 0 min, 60 min and 120 minutes after the start of digestion, C) of cacao powder.

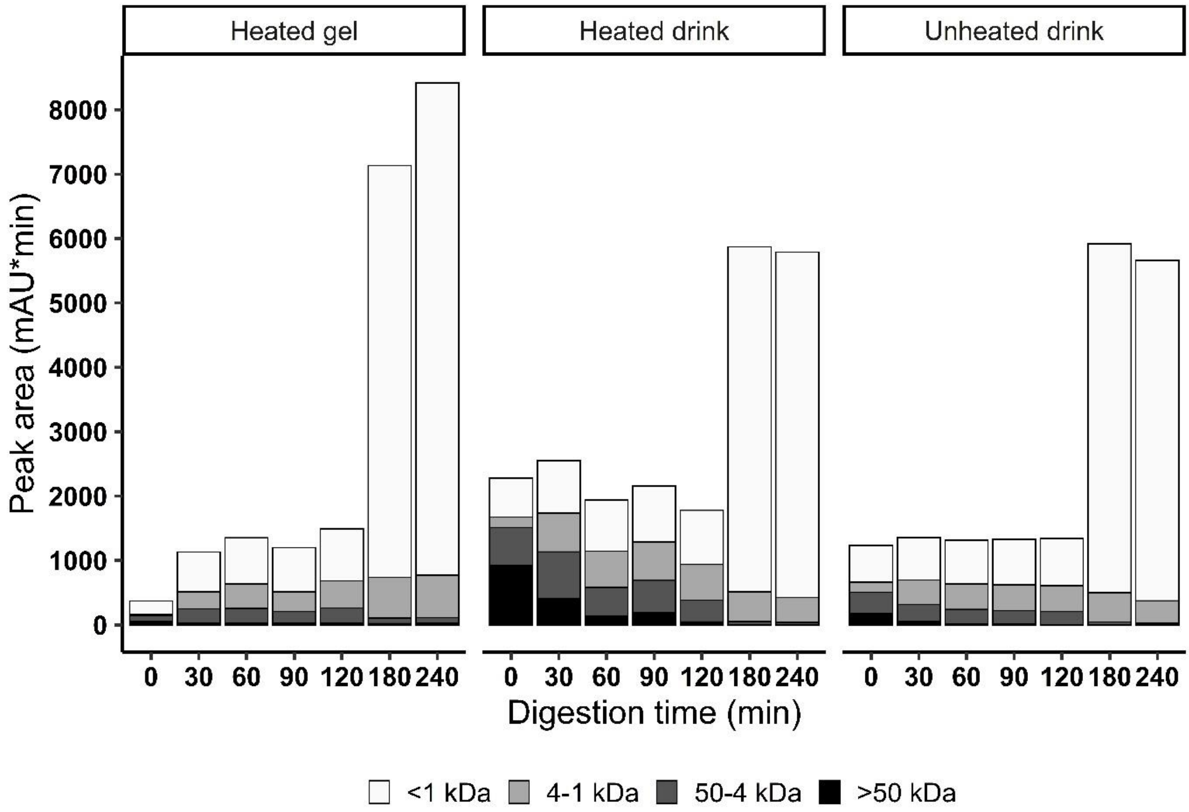


Supplementary Figure 14. Size distribution of the peptides over time (gastric phase: 0 – 120 minutes, intestinal phase: 180 and 240 minutes)
